# Prediction of Adolescent Internalizing Disorder Risk: Evidence from the Norwegian Mother, Father, and Child Cohort Study

**DOI:** 10.1101/2025.11.12.25340071

**Authors:** Evgeniia Frei, Oleksandr Frei, Espen Hagen, Alexey A. Shadrin, Nora R. Bakken, Viktoria Birkenæs, Helga Ask, Ole A. Andreassen, Olav B. Smeland

## Abstract

**Background:** Internalizing disorders are among the most common psychiatric conditions in adolescence, often associated with long-term adverse outcomes. Early identification of at-risk youth is important for effective intervention, though it remains challenging due to the multifactorial nature of risk. Machine learning (ML) offers opportunities to integrate multiple data sources and improve risk prediction for internalizing disorders.

**Methods:** We used data from 13,743 adolescents (mean age 14.45 years; 52.7% female) participating in the Norwegian Mother, Father and Child Cohort Study (MoBa), linked to national health registries. Logistic regression with elastic net regularization was applied to predict the risk of an internalizing disorder (mood, anxiety or stress-related) occurring within one to five years after assessment. Nested models of increasing complexity incorporated sociodemographic, clinical, lifestyle, mental health, psychosocial, and genetic predictors. Model performance was evaluated in a hold-out test set. Simplified models combining three questionnaire scales were also evaluated.

**Results:** Test-set performance increased with model complexity, reaching area under the receiver operating characteristic curve (AUC) of 0.732 for the full model. Mental health self-reported symptoms and psychosocial predictors contributed most to the discrimination. Simplified models using three questionnaire scales, alongside age and sex, achieved AUCs up to 0.715 and effectively stratified adolescents into high- and low-risk groups (OR80/20 ranged 6.39-10.60).

**Conclusion:** Multimodal ML models integrating registry information, mental health symptoms, psychosocial factors, and genetic data demonstrated moderate predictive performance. Simplified models with three questionnaire items reached comparable performance, highlighting their potential utility in the early identification of adolescents at elevated internalizing disorder risk.

## Introduction

Internalizing disorders are among the most prevalent disorders in adolescent psychiatry (Sacco et al., 2024), encompassing depression, anxiety, and stress-related conditions. Although symptom severity and duration vary, these disorders often entail substantial personal suffering and long-term consequences (Clayborne et al., 2019; Pollard et al., 2023), as well as a considerable burden on the healthcare system (König et al., 2020; Pollard et al., 2023).

Early identification of adolescents at elevated risk for internalizing disorders is essential for timely intervention, as targeted prevention and early support before clinical onset are likely to improve long-term outcomes (Fusar-Poli et al., 2021; Uhlhaas et al., 2023). Multiple factors spanning subclinical symptoms, environmental influences, psychological characteristics, lifestyle behaviours, and genetic determinants have been linked to internalizing disorders (Fullana et al., 2020; Köhler et al., 2018). However, a significant association between a disease and a feature does not necessarily mean that it can be used effectively for disease prediction (Bzdok et al., 2021). Current evidence suggests that the risk of internalizing disorders is not attributable to a few strong predictors, but rather to the cumulative influence of many factors, each exerting a small effect (Uher and Zwicker, 2017). Taken together, the heterogeneity and complexity of features contributing to mental illness in adolescence underscore the need for new approaches in identifying at-risk individuals (Posner, 2018; Smeland et al., 2025).

In the last decade, there has been a growing interest in applying data-driven machine learning (ML) algorithms to predict youth mental illness (Islam et al., 2024). ML offers important advantages, including the ability to handle large numbers of weak and correlated predictors, integrate multiple data modalities, and capture complex relationships (Poldrack et al., 2020). Beyond improving prediction, ML provides a useful bridge between traditional group-level association studies and clinically relevant individual-level risk estimation (Cao et al., 2024; Liu et al., 2022). Previously, population-level registries have been used to predict mental illness (Garcia-Argibay et al., 2023; Haque et al., 2021; Huang et al., 2022), but they are typically limited in terms of data modalities. Large-scale cohort studies with multimodal data overcome this limitation. Recently, ML models applied in the Adolescent Brain Cognitive Development (ABCD) and Healthy Brain Network cohorts have been used to predict adolescent mental health difficulties (de Lacy et al., 2023; Hill et al., 2025; Hou et al., 2025).

Moreover, ML framework was also used to predict adolescent depression in the Avon Longitudinal Study of Parents and Children (ALSPAC) (Yoo et al., 2024), as well as trajectories of depression symptoms in the ABCD study (Xiang et al., 2022).

Although recent cohort-based studies have advanced the field, they also have certain limitations and differ in important ways from our approach. For example, analyses based on the Healthy Brain Network cohort (de Lacy et al., 2023) relied on cross-sectional data, whereas those conducted in the ABCD study (Hill et al., 2025; Hou et al., 2025; Xiang et al., 2022) used a relatively short one- to two-year prediction horizon, focused on younger participants rather than middle adolescents, and included fewer individual measures of mental health and psychosocial functioning. Prediction modelling in the ALSPAC study (Yoo et al., 2024) utilized prenatal and childhood features, whereas our study fully leverages the depth of data collected closer to the onset of depression. Finally, the inclusion of validated diagnostic outcomes represents an important distinction of our work. The Norwegian Mother, Father, and Child Cohort Study (MoBa) (Brandlistuen et al., 2025; Magnus et al., 2016) links cohort data with clinical diagnoses from national registries, thereby combining the strengths of both registry-based clinically validated outcomes and questionnaire-based measures. Moreover, MoBa includes genetic data, offering the opportunity to test whether polygenic scores (PGS) provide additional predictive value (Kullo, 2025). Internalizing disorders show considerable heritability (Kendall et al., 2021; Ohi et al., 2025), and PGS represents a measure of innate susceptibility captured by common genetic variants. Additionally, recent studies pointed to associations between mental health and emerging environmental variables such as increasingly common use of digital devices. For example, in a recent work from our group (Frei et al., 2025) we observed significant associations between screen-based behaviors, self-reported internalizing symptoms, and registry-based psychiatric diagnoses in adolescence. Additionally, total weekend screen time was identified as an informative predictor of depressive symptoms in the ABCD study (Ho et al., 2022), and gaming time contributed significantly to prediction of mental health outcomes in vulnerable youth (Pujadas et al., 2025). Building on these findings, we included screen behaviors, specifically the time spent on social media use, gaming, and television watching, as additional candidate features for our models.

The current study had three aims. First, we evaluated how nested models of increasing complexity predict the risk of an internalizing disorder diagnosis in adolescents. Second, we assessed how much each feature set contribute to prediction. Third, we examined how to simplify the full model while maintaining adequate predictive power, to indicate which model could potentially serve as a practical tool.

## Methods

### Study sample

MoBa is a population-based pregnancy cohort study conducted by the Norwegian Institute of Public Health (Brandlistuen et al., 2025; Magnus et al., 2016). Participants were recruited from all over Norway from 1999-2008. The women consented to participation in 41% of the pregnancies. The cohort includes approximately 114,500 children, 95,200 mothers, and 75,200 fathers. Blood samples obtained from the children’s umbilical cords at birth were used for genotyping (Paltiel et al., 2014). The initial study sample included adolescents aged 14-16 years who completed the MoBa questionnaire Q-14year (response rate approximately 22%), with data linked to the Medical Birth Registry of Norway (MBRN) (*n* = 25,069). The dataset for the main ML task was restricted to participants with data available for PGS calculation after standard quality control, including exclusion of related individuals, and full questionnaire data, leaving a total of 13,743 participants. The MBRN is a national birth registry containing information about all births in Norway. More details about the study sample flow are provided in Supplementary Figure S1.

The establishment of MoBa and initial data collection was based on a license from the Norwegian Data Protection Agency and approval from The Regional Committees for Medical and Health Research Ethics. The MoBa cohort is currently regulated by the Norwegian Health Registry Act. The current study was approved by The Regional Committees for Medical and Health Research Ethics (2016/1226/REK Sør-Øst C) in Norway.

### Outcome variables

Information about psychiatric diagnoses in the study sample was retrieved from the Norwegian Patient Registry (NPR; diagnostic information 2008-2024), as well as the Norwegian Control and Payment of Health Reimbursements Database (KUHR) and the Norwegian Registry for Primary Health Care (NRPHC; diagnostic information 2006-2024). The primary outcome was the presence of an internalizing disorder diagnosis, defined as having at least one relevant code recorded in either the NPR (ICD-10) (World Health Organization, 2019) or the KUHR/NRPHC (ICPC-2) (Hofmans-Okkes and Lamberts, 1996). Relevant ICD-10 codes included F32-F34 (depressive disorders and persistent mood [affective] disorders); F38 (other affective disorders); F39 (unspecified affective disorders); F40-F43, F93.0, F93.1, F93.2 (anxiety and stress-related disorders). Relevant ICPC-2 codes included P79 (phobia/compulsive disorder), P76 (depressive disorder), P74 (anxiety disorder), and P82 (post-traumatic stress disorder). To define a prediction window, only diagnoses occurring between one and five years after completion of the Q-14year questionnaire were included. Participants who received an outcome diagnosis before the 1-year cutoff were excluded to prevent label leakage (see Figure S1).

### Predictor variables

Age (at the Q-14year questionnaire completion) and sex assigned at birth were included as baseline predictors in all models. Sociodemographic factors included immigrant status, highest parental education, and household income. Those variables were obtained from national registry data provided by Statistics Norway (SSB) (see Supplementary Text 1). Clinical and family history predictors comprised relevant diagnostic codes from national health registries. Specifically, diagnoses F92 (Mixed disorders of conduct and emotions), F93 (Emotional disorders with onset specific to childhood), F94 (Disorders of social functioning with onset specific to childhood and adolescence), F95 (Tic disorders), or F98 (Other behavioural and emotional disorders with onset usually occurring in childhood and adolescence) received prior to questionnaire completion, neurodevelopmental disorder diagnoses (F90 [Hyperkinetic disorders] and F84 [Pervasive developmental disorders]) and other psychiatric diagnoses (i.e., ICD-10 F-codes not specified above and not included as outcome variables), were included as separate model features. ICPC-2 codes P01 (Feeling anxious/nervous/tense), P02 (Acute stress reaction), and P03 (Feeling depressed), which represent non-specific psychological complaints recorded in primary care, were also used as predictors. Additionally, we included parental internalizing disorder diagnoses.

Information on lifestyle factors (sleep, alcohol or drug use, smoking, screen behaviors, exercise) was retrieved from the Q-14year questionnaire. Adolescent self-reported mental health symptoms and psychosocial functioning were assessed in the same questionnaire using several instruments (see Table 1, Table S1). Items were reverse coded where necessary so that high scores reflected greater symptom load. Detailed information about all questionnaire items is presented in the supplement (Table S1). Analyses included only participants with complete data.

**Table 1.** Overview of predictor groups used in ML models.

| **Predictor** | **Source** | **N questions** |
| --- | --- | --- |
| *Baseline predictors* |  |  |
| Age when the questionnaire was filled out | Q14-year | NA |
| Sex assigned at birth | MBRN | NA |
| *1. Sociodemographic factors* |  |  |
| Immigrant status | SSB | NA |
| Parental education | SSB | NA |
| Household income | SSB | NA |
| *2. Clinical and family history* |  |  |
| Neurodevelopmental diagnosis | NPR | NA |
| Other childhood-onset psychiatric disorder diagnosis | NPR | NA |
| Other ICD-10 F-diagnosis | NPR | NA |
| Parental internalizing disorder diagnosis | NPR | NA |
| Primary care psychological symptom codes (ICPC-2) | KUHR/<br>NRPHC | NA |
| *3. Lifestyle factors* |  |  |
| Questions about sleeping problems | Q14-year | 3 |
| Questions about experience with alcohol and use of drugs | Q14-year | 4 |
| Questions about the child's smoking/snusing habits | Q14-year | 5 |
| Time spent watching TV | Q14-year | 1 |
| Time spent gaming | Q14-year | 1 |
| Time spent communicating with friends on social media | Q14-year | 1 |
| Exercise, days per week | Q14-year | 1 |
| *4. Mental health symptoms* |  |  |
| Short Mood and Feelings Questionnaire (SMFQ) | Q14-year | 13 |
| The (Hopkins) Symptoms Checklist (SCL-10) | Q14-year | 10 |
| Mini Social Phobia Inventory (MiniSPIN) | Q14-year | 3 |
| Screen for Child Anxiety Related Disorders (SCARED) | Q14-year | 5 |
| Eating Disorder Examination Questionnaire (EDE-Q) (selected questions*) | Q14-year | 8 |
| Parent/Teacher Rating Scale for Disruptive Behaviour Disorders (RS-DBD) (selected questions) | Q14-year | 8 |
| *5. Psychosocial domain* |  |  |
| Self-Perception Profile for Adolescents, Scale for Social Competence (adapted questions) | Q14-year | 5 |
| Strengths and Difficulties Questionnaire, Prosocial Subscale | Q14-year | 5 |
| Satisfaction with Life Scale | Q14-year | 5 |
| Differential Emotional Scale (DES), Enjoyment Subscale | Q14-year | 3 |
| Parental relations Self-concept Scale from the Self-Description Questionnaire II-Short (SDQII-S) (selected questions) | Q14-year | 4 |
| Parent-Child Conflict Scale; the Parental Environment Questionnaire (selected questions) | Q14-year | 4 |
| Questions concerning being bullied | Q14-year | 4 |
| Rosenberg Self-Esteem Scale (selected questions) | Q14-year | 4 |
| IPIP Big-Five factor markers (selected questions): extraversion | Q14-year | 4 |
| IPIP Big-Five factor markers (selected questions): agreeableness | Q14-year | 4 |
| IPIP Big-Five factor markers (selected questions): conscientiousness | Q14-year | 4 |
| IPIP Big-Five factor markers (selected questions): emotional stability | Q14-year | 4 |
| IPIP Big-Five factor markers (selected questions): intellect | Q14-year | 4 |
| Serious Life Event | Q14-year | 6 |
| *6. Polygenic Scores* |  |  |
| Major depression PGS | NA | NA |
| Anxiety PGS | NA | NA |
| Bipolar disorder PGS | NA | NA |

MBRN: Medical Birth Registry of Norway; NPR: Norwegian Patient Registry; KUHR: Norwegian Control and Payment of Health Reimbursements Database; NRPHC: Norwegian Registry for Primary Health Care; SSB: Statistics Norway; NA: not applicable; IPIP: the International Personality Item Pool; PGS: polygenic score.
\* "Selected questions" indicate that a subset of items from the full instrument was selected to use in the MoBa study.

### Genetic data

Full details about the genotyping and QC procedure are provided elsewhere (Corfield et al., 2024). To estimate PGS of psychiatric disorders we used summary statistics from recent large-scale GWASs of major depression (MD), anxiety (ANX), and post-traumatic stress disorder (PTSD) (Friligkou et al., 2024; Levey et al., 2021; Nievergelt et al., 2024). PGS were calculated using SBayesR (Lloyd-Jones et al., 2019) as implemented in the reference-standardized, reproducible GenoPred pipeline (Pain et al., 2024). The sample for PGS analysis was restricted to unrelated participants of European-like ancestry. More details are provided in the Supplementary Text 2.

### Machine learning (ML) approach

The dataset (*n =* 13,743) was split into training (80%) and hold-out test (20%) sets using a stratified split to maintain the same proportion of cases. On the training set, we fitted logistic regression models with elastic net regularization (Zou and Hastie, 2005), and mixing parameter (α = 0.5) using *glmnet* (R). In *glmnet*, predictors were internally standardized to a mean of zero and scaled to unit variance before model fitting; the same scaling was automatically applied to the test data. The hyperparameter (λ) was tuned using a nested 10-fold cross-validation within the training set, aiming to maximize area under the receiver operating characteristic curve (AUC) (see Supplementary Text 3). The final model was refit on the full training set and evaluated on the hold-out test set. To assess robustness to the choice of the mixing parameter, additional models were trained with α = 0, 0.25, 0.75, and 1. Model performance was primarily evaluated on the hold-out test set using AUC with 95% confidence intervals (CIs) obtained by 1,000 bootstrap resamples.

After fitting the model, per-subjects risk scores were computed on the test data, with higher scores linked to the higher probability of receiving a diagnosis. Using those risk scores, the test-set participants were stratified into low (≤20^th^ percentile), medium (21–79^th^ percentile), and high (≥80^th^ percentile) risk groups, and odds ratios (ORs) for internalizing disorders were calculated by comparing high-risk and low-risk groups (OR80/20). Sensitivity, specificity, balanced accuracy, positive predictive value (PPV), negative predictive value (NPV), and confusion matrices were estimated in the test set using the optimal threshold determined by Youden’s index (Youden, 1950) calculated in the training set. In addition, we assessed performance at a threshold corresponding to 50% sensitivity.

### Nested models with increased complexity

We built a sequence of models with progressively larger feature sets, with all models including age and sex as baseline covariates:

- Model 1: sociodemographic factors (5 predictors)
- Model 2: model 1 + clinical and family history (10 predictors)
- Model 3: model 2 + lifestyle factors (17 predictors)
- Model 4: model 3 + mental health symptoms (23 predictors)
- Model 5: model 4 + psychosocial domain (33 predictors)
- Model 6 (full model): model 5 + PGSs (36 predictors).

An overview of predictor groups is presented in Table 1. Each predictor group was also evaluated in separate models.

To confirm that performance exceeded chance levels, we applied permutation testing (1000 permutations) to the full model. In each permutation, outcome labels were randomly shuffled, and the full modelling procedure was repeated to generate a null distribution of AUC values. As an additional sensitivity analysis, we performed leave-one-region-out validation, treating each of the four Norwegian health authority regions as a hold-out test set to assess the generalizability of the full model to independent sites and participants.

### Simplified (triplet) models

We additionally evaluated whether a small subset of questionnaire items, added to age and sex, could yield adequate predictive performance. For this analysis, we used an extended training set (*n* = 14,716; participants with full questionnaire data, not restricted to genetic data), while the test set was identical to that used for the nested models (see Supplementary Text 4). Single predictors from three domains – lifestyle factors, mental health symptoms, psychosocial functioning – were first screened in the extended training set, and the top 10 with the highest AUC values were retained. For each retained predictor, we identified the most informative additional predictor that gave an improvement of at least ΔAUC ≥ 0.02. From each such pair we created triplets by adding one further predictor and estimated their training performance. Triplets for test-set evaluation were then selected using a greedy set-packing procedure that prioritized higher AUC values (Cormen et al., 2009). Each questionnaire scale was initially restricted to a single use, with the constraint relaxed as needed (i.e., if fewer than the target number of triplets could be selected) to allow at most two uses per scale. The maximum target number of triplets was set to 10.

## Results

Of the 25,069 adolescents who completed the Q-14year questionnaire and had MBRN data, 2,155 had received an internalizing disorder diagnosis before the prediction window and were excluded from analysis. Among the remaining participants, 13,743 had both genetic data and complete questionnaire data (Figure S1). Those comprised the final analytic sample for the nested model analysis (mean age 14.45 years; 52.69% female). Within this sample, 1,004 (7.31%) received an internalizing disorder diagnosis during the prediction window. The sample was divided into a training set (80%, n = 10,994) and a hold-out test set (20%, n = 2,749), maintaining the same proportion of cases. Main sociodemographic characteristics of the overall sample, training and test sets are shown in Table 2. Full descriptive statistics for all predictors are provided in Table S2.

**Table 2.** Sociodemographic characteristics of the full sample, the training set, and the hold-out test set.

| Variable | Full sample<br>(*n* = 13,743) | Training set<br>(*n* = 10,994) | Test set<br>(*n* = 2,749) |
| --- | --- | --- | --- |
| Age in years, mean (SD) | 14.45 (0.52) | 14.44 (0.52) | 14.46 (0.52) |
| Female sex assigned at birth, N (%) | 7,242 (52.70%) | 5,782 (52.59%) | 1,460 (53.11%) |
| Household income low, N (%) | 197 (1.43%) | 157 (1.43%) | 40 (1.46%) |
| Parental education low, N (%) | 2,199 (16.00%) | 1,752 (15.94%) | 447 (16.26%) |
| Immigrant status, N (%) | 110 (0.80%) | 85 (0.77%) | 25 (0.91%) |
| Internalizing disorder diagnosis, N (%) | 1,004 (7.31%) | 803 (7.30%) | 201 (7.31%) |

### Nested models

The performance of the nested models is summarized in Figure 1, Table 3 and Figure S2. The baseline model, including only sociodemographic factors, achieved an AUC of 0.626 (95% CI: 0.590-0.665) in the test set. Performance increased steadily with the addition of features, reaching 0.732 (95% CI: 0.696-0.766) for the full model with all predictor groups. When evaluated in separate models (with age and sex), self-reported mental health symptoms and psychosocial predictors showed the highest discriminative performance, with test AUC 0.710 (95% CI: 0.671-0.743) and 0.703 (95% CI: 0.664-0.737), respectively. Other predictor groups demonstrated more modest performance, with test AUC ranged 0.610-0.699 (Table S4, Figure S3).

**Figure 1.**
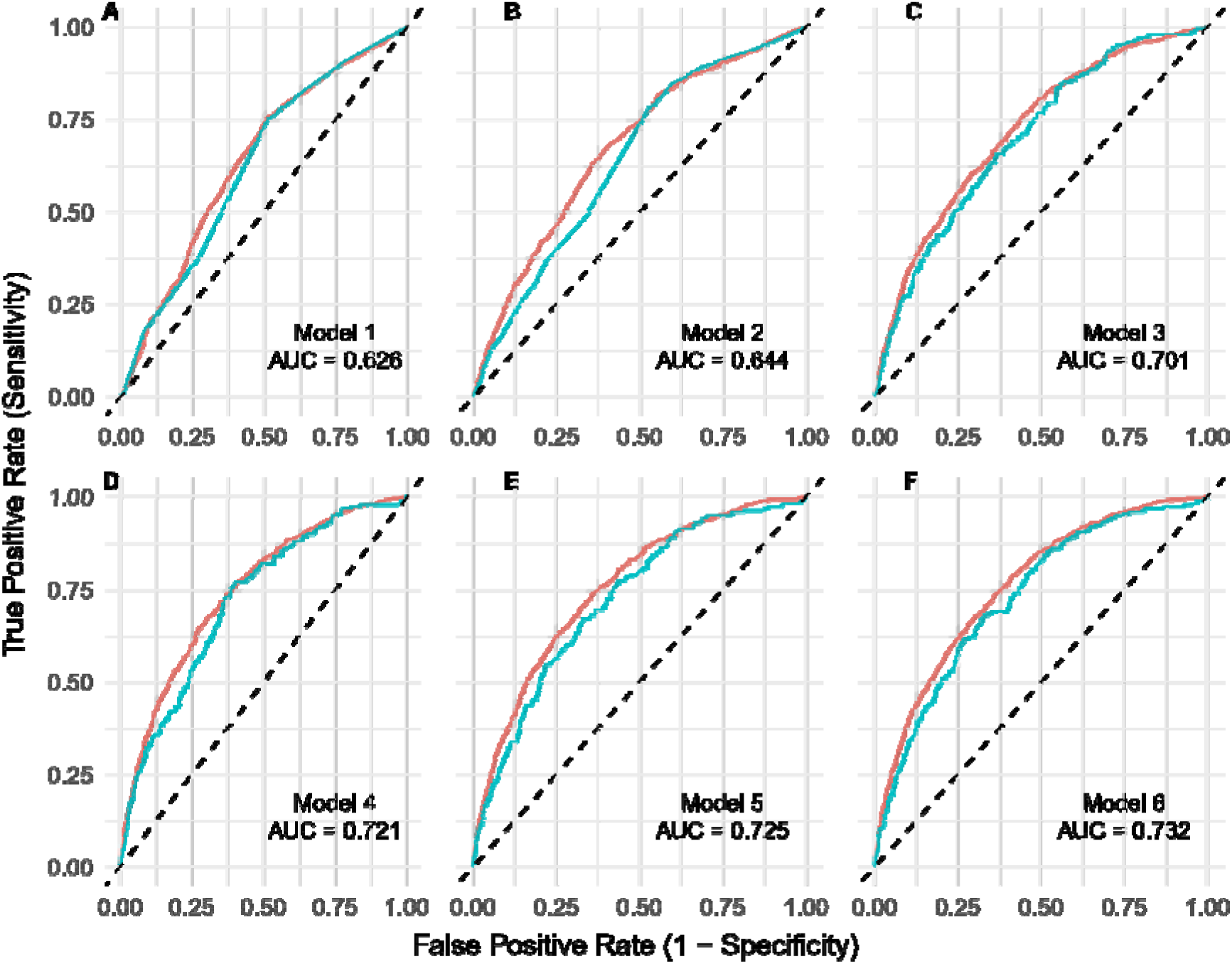
Discriminative performance (AUC) of nested models in training and independent test sets. Subplots (A-F) show the performance of six nested models, each evaluated in the training set (red line, 95% CI shown as grey band) and in the hold-out test set (blue line). AUC: area under the receiver operating characteristic (ROC) curve. Confidence bands around ROC curves represent pointwise 95% CIs for sensitivity at fixed specificities, as implemented in the pROC package. Model 1: sociodemographic factors; Model 2: model 1 + clinical and family history; Model 3: model 2 + lifestyle factors; Model 4: model 3 + mental health symptoms; Model 5: model 4 + psychosocial domain; Model 6 (full model): model 5 + polygenic scores.

**Table 3.** Performance of the nested models for predicting internalizing disorders.

|  | Predictors | AUC test (95% CI) | OR80/20 (95% CI) |
| --- | --- | --- | --- |
| Model 1 | Sociodemographic factors | 0.626 (0.590-0.665) | 3.42 (1.96-5.97) |
| Model 2 | Model 1 + clinical and family history | 0.644 (0.609-0.682) | 5.15 (3.03-8.46) |
| Model 3 | Model 2 + lifestyle factors | 0.701 (0.665-0.735) | 18.06 (7.36-48.00) |
| Model 4 | Model 3 + mental health symptoms | 0.721 (0.686-0.755) | 19.77 (8.00-50.68) |
| Model 5 | Model 4 + psychosocial domain | 0.724 (0.690-0.756) | 13.85 (6.68-30.35) |
| Model 6 | Model 5 + polygenic scores | 0.732 (0.696-0.766) | 17.02 (7.41-38.94) |

In the test set, the full model (Model 6) achieved a sensitivity of 0.632 (95% CI 0.562-0.701), specificity of 0.701 (95% CI 0.682-0.717), balanced accuracy of 0.666 (95% CI 0.629-0.702), PPV of 0.143 (95% CI 0.128-0.157), and NPV of 0.960 (95% CI 0.953-0.968) at the optimal threshold determined by Youden’s index. Performance metrics and confusion matrices for all nested models are presented in Table S5, and Figures S4, S5.

Permutation testing confirmed that the full model performed significantly above chance (p < 0.001). In the leave-one-region-out analysis, test AUCs indicated consistent generalizability of the full model to independent sites (Table S6). Sensitivity analyses across different α values (0, 0.25, 0.75, 1) demonstrated stable performance of the full model regardless of the mixing parameter (Table S7). OR80/20 values increased substantially from the simplest to the more complex models, indicating that additional predictors improved risk stratification. Odds of internalizing disorders increased progressively across risk strata, with the medium group showing 2.61- to 7.67-fold higher odds than the lowest group, and the highest group showing 1.64- to 3.93-fold higher odds than the rest of the sample (Table S3).

### Simplified (triplet) models

During training, the best performing pairs of predictors (together with age and sex) achieved AUC up to 0.724 (Figure S6). Among the 436 candidate triplet models, all showed improvements over their corresponding pairs in training, with AUC ranging from 0.697 to 0.727 (Figures S7, S8). The greedy set-packing procedure selected 7 high-performing, low-redundancy triplets out of a maximum 10; these were evaluated on the test set (Table 4).

**Table 4.** Performance of simplified (triplet) models predicting internalizing disorder diagnosis, selected based on training set performance.

|  | **Predictors** | **AUC test**<br>**(95% CI)** | **OR80/20** | **N questions** |
| --- | --- | --- | --- | --- |
| Triplet 1 | Mental distress + Sleep + Exercise | 0.715<br>(0.676-0.750) | 9.36<br>(4.68-18.55) | 14 (10 + 3 + 1) |
| Triplet 2 | Social competence + Emotional stability + Life satisfaction | 0.700<br>(0.659-0.736) | 8.91<br>(4.78-17.27) | 14 (5 + 4 + 5) |
| Triplet 3 | Anxiety symptoms + Self-esteem + Enjoyment | 0.695<br>(0.659-0.731) | 8.41<br>(4.54-16.70) | 12 (5 + 4 + 3) |
| Triplet 4 | Eating problems + Mental distress + Exercise | 0.705<br>(0.668-0.741) | 7.44<br>(4.05-13.61) | 19 (8 + 10 + 1) |
| Triplet 5 | Anxiety symptoms + Life satisfaction + Emotional stability | 0.700<br>(0.662-0.736) | 6.39<br>(3.67-11.17) | 15 (5 + 5 + 5) |
| Triplet 6 | Sleep + Depressive symptoms + Social competence | 0.715<br>(0.680-0.750) | 10.60<br>(5.41-20.24) | 21 (3 + 13 + 5) |
| Triplet 7 | Eating problems + Enjoyment + Depressive symptoms | 0.693<br>(0.655-0.730) | 7.04<br>(3.87-12.97) | 24 (8 + 3 + 13) |

AUC: Area under the receiver operating characteristic curve; Mental distress: SCL-10, The Hopkins Symptoms Checklist; Social competence: Self-Perception Profile for Adolescents, Scale for Social Competence (adapted questions); Life satisfaction: SWLC, Satisfaction with Life Scale; Emotional stability: IPIP\_stability, International Personality Item Pool Big-Five factor markers, selective items related to emotional stability factor; Anxiety symptoms: SCARED, Screen for Child Anxiety Related Disorders; Self-esteem: RSES, Rosenberg Self-Esteem Scale (selected questions); Eating problems: EDE-Q, Eating Disorder Examination Questionnaire (selected questions); Enjoyment: DES, Differential Emotional Scale, Enjoyment Subscale; Depressive symptoms: SMFQ, Short Mood and Feelings Questionnaire.
95% confidence intervals (CIs) obtained by 1,000 bootstrap resamples.

## Discussion

Here we utilized MoBa data to develop ML models for predicting adolescents’ internalizing disorder risk. The full model – comprising all candidate features across domains – achieved an AUC of 0.732. Though all nested models were able to stratify participants on internalizing disorder risk, the performance improved stepwise, with strongest contributions from mental health symptom scales and psychosocial predictors. Additionally, we evaluated seven simplified models that may be suitable for practical implementation, as they achieved predictive performance comparable to the more complex ones.

To interpret these results, it is important to note that while some sources describe AUC values between 0.7 and 0.8 as “acceptable” or “fair”, no universal terminology exists (White et al., 2023), and some authors recommend reporting AUC values without categorical labels (de Hond et al., 2022). Therefore, we intended to use AUC values mainly to compare the discriminative ability of different models. Beyond discrimination, however, performance metrics provide a complementary perspective on practical utility. Our models showed low PPV but high NPV, a pattern consistent with the relatively low base rate of adolescent internalizing disorders. Nonetheless, the top-risk stratum represented a marked enrichment of true cases. In practical terms, the models are best viewed as tools for population-level risk stratification rather than individual-level diagnostic classification, consistent with their intended purpose.

When evaluating nested models, stepwise improvements in performance was accompanied by increasing OR80/20 values. However, the largest odds ratios – seen in the most complex models – had the widest confidence intervals, reflecting less precise estimates due to fewer events in extreme groups. We also observed that even simple predictor sets could stratify participants by internalizing disorder risk. For instance, registry-only sociodemographic and clinical history data without additional questionnaire items (Model 3) provided meaningful risk stratification, consistent with prior findings demonstrating the predictive utility of health records for diverse mental health outcomes (Garriga et al., 2022; Raket et al., 2020; Su et al., 2020). PGS contributed little to model performance once comprehensive phenotypic data were included, in line with earlier studies (Agerbo et al., 2021; Høberg et al., 2024; Lu et al., 2023). Nevertheless, in a parsimonious model including age, sex, and PGS, performance exceeded the null model (age and sex only) by ΔAUC = 0.055 (95% CI: 0.026-0.080). This relatively small but statistically significant improvement may be practically useful due to the low incremental burden and any-time availability of PGS when genetic data are already collected (Lambert et al., 2019). Mental health symptoms and psychosocial predictors contributed most to the performance of the nested models and, when evaluated separately alongside age and sex, were the best-performing predictor sets. This aligns with prior evidence on the predictive utility of questionnaire data (Hill et al., 2025); importantly, we demonstrate it in a registry-linked cohort with verified clinical outcomes and focusing solely on youth perspective, in contrast to studies that also relied on parental reports (Hill et al., 2025; Ho et al., 2022).

While complex multimodal models may provide a gain in predictive performance over unimodal models (Soenksen et al., 2022), they are still difficult to implement in clinical practice. We therefore selected and evaluated seven simpler models that are more amenable to translation into real-world settings. Each of these models, comprising three questionnaire scales from the lifestyle, mental health, and psychosocial domains, as well as age and sex, showed potential for relevant risk stratification. Odds of internalizing disorders increased monotonically across predicted risk strata, with the strongest contrast observed between the high- and low-risk groups (OR80/20 ranged 6.39-10.60), and a clear separation of the high-risk group from the remainder of the sample. This underscores the potential of those parsimonious models to identify individuals at particularly elevated risk in applied settings. Importantly, the performance of the simplified three-feature models was comparable to that of models including all mental health symptom scales (five features in total) or all psychosocial predictors (ten features in total) evaluated as separate sets. Interestingly, the questionnaire item describing exercise did not rank among the top individual predictors, but it consistently enhanced the performance of predictor pairs and was included in the selected triplet models. It stood apart from other lifestyle factors such as smoking, alcohol use, and screen time, and given its simplicity as a single-item measure, it may have practical utility. Screen use measures did not emerge as strong predictors in our models, suggesting that while screen behaviours are associated with adolescent mental health (Frei et al., 2025), their contribution to risk stratification is modest when considered alongside psychosocial and symptom-based measures, particularly when the outcome is clinically diagnosed internalizing disorders. Our findings also support the role of sleep quality as a potentially important predictor of internalizing disorder risk, consistent with prior studies (Bacaro et al., 2023; Ho et al., 2022).

Despite these informative findings, some limitations should be considered. First, the absence of a truly independent test set (i.e., external validation) makes it difficult to fully assess the generalizability of the models. Although the leave-one-region-out sensitivity analysis supported generalizability across health authority regions, non-representativeness and selection bias – which are well-documented challenges in cohort studies (Biele et al., 2019; Nilsen et al., 2009; Vejrup et al., 2022) – could limit generalizability in other populations. Second, because the MoBa cohort spans more than a decade and includes participants recruited at different birth years, follow-up times inevitably vary. As a result, not all participants had the full prediction window covered, which may have led to an underestimation of the true sensitivity of our models. The maximum AUC in our study was somewhat lower than in previous analyses of the ABCD cohort based on both parent and youth reports (Hill et al., 2025), where predictive performance was driven largely by parent-reported measures – unlike our models, which relied on youth self-reports. Moreover, many MoBa instruments were administered in shortened versions or with only selected questions. Using full versions of these instruments might improve prediction – particularly for constructs previously found to be important, such as family environment (Ho et al., 2022) – but would also increase model complexity. The same applies to longitudinal aspects (ongoing behavioural assessments, dynamic environmental influences, sleep trajectories, etc.) which could further enhance model performance but at the cost of markedly greater model sophistication. The trade-off between predictive performance and feasibility needs careful consideration – an issue we explicitly addressed in this study. Finally, we excluded participants with early diagnoses of internalizing disorders to prevent label leakage. While this was a necessary step for model development, it may have introduced some imbalance in terms that individuals with early-diagnosed disorders – who might display more severe or persistent symptoms – were not analysed. This restriction could therefore limit the generalizability of the models to the full spectrum of the internalizing pathology.

In conclusion, we investigated a wide range of features (sociodemographic and registry data, lifestyle factors, mental health scales, psychosocial predictors, genetics) and demonstrated stepwise improvement of performance, with the full model reaching AUC 0.732. We replicated several previous findings in a large cohort with clinically confirmed diagnoses, narrowing the feature space for future prediction studies. Finally, our findings show that a small set of questionnaire scales can achieve discrimination comparable to complex feature sets. Together, these findings can form the basis for development of better early identification of high-risk youth who may benefit from additional support to improve mental health outcomes.

### Data availability

Data from the Norwegian Mother, Father and Child Cohort Study is managed by the Norwegian Institute of Public Health. Access requires approval from the Regional Committees for Medical and Health Research Ethics (REC), compliance with GDPR, and data owner approval. Participant consent does not allow individual-level data storage in repositories or journals. Researchers seeking access for replication must apply via www.helsedata.no.

## Funding sources

This work was supported by the Research Council of Norway (grant number 324499, awarded to Ole A. Andreassen).

## Acknowledgements

We are grateful to all the participating families in Norway who take part in this on-going cohort study. For generating high-quality genomic data, we thank the Norwegian Institute of Public Health (NIPH), the HARVEST collaboration, the NORMENT Centre at the University of Oslo, the Center for Diabetes Research at the University of Bergen, deCODE Genetics, the Research Council of Norway, the South-Eastern and Western Norway Regional Health Authorities, the ERC AdG, Stiftelsen KG Jebsen, the Trond Mohn Foundation, and the Novo Nordisk Foundation. This work was performed on Services for sensitive data (TSD), University of Oslo, Norway, with resources provided by UNINETT Sigma2 - the National Infrastructure for High Performance Computing and Data Storage in Norway.

## Declaration of generative AI use and AI-assisted technologies in the manuscript preparation process

During the preparation of this work the author(s) used GPT UiO and ChatGPT-5 to review parts of the code, improve text clarity and readability (for example, by suggesting synonyms to reduce redundancy), and check grammar and spelling. After using these tools, the author(s) reviewed and edited the content as needed and take(s) full responsibility for the final version of the manuscript.

## Declaration of interest

Professor Ole A. Andreassen has received speaker fees from Lundbeck, Janssen, Otsuka, and Sunovion, and is a consultant to Cortechs.ai, and Precision Health AS. Dr. Oleksandr Frei is a consultant to Precision Health AS. Dr. Evgeniia Frei and Dr. Oleksandr Frei are spouses. No potential conflict of interest was reported by other authors.

## Notes

### Author Declarations

The establishment of MoBa and initial data collection was based on a license from the Norwegian Data Protection Agency and approval from The Regional Committees for Medical and Health Research Ethics. The MoBa cohort is currently regulated by the Norwegian Health Registry Act. The current study was approved by The Regional Committees for Medical and Health Research Ethics (2016/1226) in Norway.

