## Supplementary text, tables, and figures for "Prediction of Adolescent Internalizing Disorder Risk: Evidence from the Norwegian Mother, Father, and Child Cohort Study"

**Supplementary Note**

### Supplementary Text

*1. Phenotypic Variables*

Immigration background was defined using national registry data provided by Statistics Norway (SSB). Participants were classified as having an *immigrant background* (coded as 1) if they were either:

- Born abroad to two foreign-born parents (SSB code B), or
- Born in Norway to two foreign-born parents (SSB code C).

Individuals with no immigrant background (SSB code A), or with partial Norwegian background (E, F, G) were classified as having *non-immigrant background* (coded as 0) for the purposes of this analysis. This approach aligns with SSB’s definitions used in national population statistics.^1^

Parental level of education was obtained from the National Education Database via SSB, using the Norwegian Standard Classification of Education (NUS2000)^2^ as the measurement framework. We extracted the three-digit NUS2000 codes, representing highest completed education at the time the child was 16 years old. This was used as a proxy for education at the time of Q-14year completion, as parental education is typically stable during adolescence. The score of the parent with the highest education level (or the one available, in case one score was missing) was chosen to represent parental educational level. Parental education was classified according to NUS2000 as mapped to ISCED 2011 levels. Low education (coded as 1) was defined as NUS2000 codes 0-5, reflecting no, primary, secondary, or post-secondary non‑tertiary education. High education (coded as 0) was defined as NUS2000 codes 6-8, corresponding to university-level education (bachelor’s degree or higher). This grouping reflects the high overall educational level in the study population and is consistent with a stricter definition of tertiary education as an indicator of socioeconomic advantage. The coding scheme reflects the role of low parental education as a risk factor for adverse mental health outcomes.^3^

Household income was defined using the registry variable that represents after-tax income per consumption unit (European Union scale). To classify low income, we compared each participant’s household income to the national median for the corresponding calendar year (i.e., the year of questionnaire completion). Individuals were coded as having low household income (coded as 1) if parental income was below 60% of the national median, in line with standard definitions from SSB.

Instruments used in the Q-14year questionnaire for assessment of lifestyle factors, mental health symptoms, and psychosocial functioning are presented in Table S1. Each of the IPIP Big Five personality factors was included as a separate predictor in the model, based on the following items: Extraversion (1, 6r, 11, 16r), Agreeableness (2, 7r, 12, 17r), Conscientiousness (3, 8r, 13, 18r), Emotional Stability (4r, 9, 14r, 19r), and Intellect (5, 10, 15r, 20r). A mean score of the items was computed for each instrument, requiring at least half the items to be non‑missing. The mean was then multiplied by the number of items in the instrument to give a representative score on the scale of the instrument, following the approach used in previous studies based on the MoBa data.^4^ Items were reverse coded where necessary so that high scores reflected greater symptom load. Participants were excluded from analysis if the mean score of any instrument could not be calculated (i.e., more than half of the items were missing).

*2. Genetic data*

To estimate polygenic scores (PGS) of psychiatric disorders we used summary statistics from recent large-scale GWASs of bipolar disorder (BD), major depression (MD), and anxiety (ANX), independent of the MoBa sample^5–7^. For PGS calculation the sample was restricted to unrelated participants of European ancestry following the GenoPred QC procedure^8^. Subjects with inconsistencies between sex assigned at birth and genotyped sex and subjects with chromosomal abnormalities (as indicated in the MBRN) were manually removed from analyses (n < 5). We used PGSs calculated with SBayesR^9^, one of the recommended methods for application in psychiatric disorders^10^.

*3. Machine learning approach*

Related individuals were excluded during PGS calculation, and verification of mother IDs confirmed that no maternal siblings were present within or across data splits. Therefore, familial relatedness was not a concern for data leakage. We fit logistic regression models with an elastic net penalty (α = 0.5; α = 0 corresponds to Ridge regression, α = 1 corresponds to Lasso regression) using the *glmnet* package in R. PGSs were pre-residualized for genotyping batch and first 10 principal components using linear models fit on the training set; the resulting coefficients were then applied to the hold-out test set to prevent information leakage. Random partitions were made reproducible with a fixed seed.

The hyperparameter λ was selected using a nested cross-validation procedure on the training set. The outer loop used 10-fold cross-validation (CV) to estimate model performance, and within each outer-training partition the inner loop used cv.glmnet (nfolds = 10) to select the regularization parameter λ that maximized the mean cross-validated area under the receiver operating characteristic (ROC) curve (AUC). The model trained on the outer-training partition was then applied to the outer-test partition to obtain out-of-fold scores; these predictions (one per sample across all outer folds) were then pooled and later used, for example, to estimate Youden’s index. After nested CV, we refit cv.glmnet on the entire training set (with nfolds = 10 to choose the final λ), and evaluated the resulting model on the hold-out test set, reporting AUC with 95% CIs obtained by 1,000 bootstrap resamples. To access the robustness of the results to the balance of L1 and L2 regularization, we repeated the full procedure for α ∈ {0, 0.25, 0.75, 1}.

Sensitivity, specificity, balanced accuracy, positive predictive value (PPV), negative predictive value (NPV), and confusion matrices for the nested models were calculated at the optimal cutoff determined by Youden’s index from the ROC curve, with 95% CIs obtained by 1,000 bootstrap resamples. Youden’s index was estimated from the ROC curve derived from pooled cross-validated predictions in the training set and then applied to the test set. The same procedure was used in sensitivity analyses (e.g. estimating performance of the full model across different alpha values).

The prediction window in our study was defined as the period between one and five years after completion of the Q-14year questionnaire. Because the MoBa cohort spans more than a decade and includes participants recruited at different birth years, follow-up times inevitably varied. Consequently, not all participants had the entire prediction window covered. While ≈35% of the sample had complete coverage of the full prediction window, ≈82% had follow-up data for at least 24 months within the prediction period.

*4. Evaluating performance of small predictor sets*

For this analysis, we used an extended training set not restricted to participants with genetic data, while the test set was identical to that used for the nested models. To avoid data leakage due to relatedness, we pruned the training set to exclude any participant sharing a mother ID with someone in the test set. That resulted in an extended training set of 14,716 individuals.

First, we evaluated the predictive performance of each score individually by fitting models of the form: F ~ sex + age + score_1, where “score_1” represents one predictor from the evaluated domains (health and lifestyle factors, mental health symptoms, psychosocial functioning). Next, to assess potential synergistic effects, we evaluated all pairwise combinations of the scores in models of the form: F ~ sex + age + score_i + score_j, for all unique pairs i ≠ j. Model performance was assessed in the extended training set using AUC.

We first retained 10 predictors with the highest training AUC values and, for each, identified the most informative additional predictor defined as one yielding an improvement of at least ΔAUC ≥ 0.02. For each such pair (*n* = 20), we systematically added one further predictor (along with age and sex as baseline covariates), resulting in 436 unique triplet models whose performance was estimated in the training set. From these, we selected 7 triplets for test-set evaluation using a greedy set-packing procedure that prioritized higher AUC values. Each questionnaire scale was initially restricted to a single use, with the constraint relaxed as needed (i.e., if fewer than the target number of triplets could be selected) to allow at most two uses per scale. The maximum target number of triplets was set to 10.

### Supplementary Tables

### Table S1. Instruments from the Q14-year questionnaire used in the study.

| **Question** | **Response options** | | **Variable name** |
| --- | --- | --- | --- |
| **Screen time use** | | | |
| How much time do you usually spend during one weekday on the following activities? | | | |
| Watch movies/series/TV | 1-Never/rarely  2-Less than 1 hour  3-1-2 hours  4-3-4 hours  5-5-6 hours  6-7 hours or more | | UB18 |
| Playing games (on PC, TV, tablet, mobile etc.) |  |  | UB20 |
| Sitting/lying down with PC, mobile or tablet (irrespective of activity) |  |  | UB21 |
| Communicating with friends on social media |  |  | UB22 |
| **Physical activity** | | | |
| Outside school hours, how many days a week do you usually do the following? | | | |
| Exercise (e.g. soccer, handball, skiing, running, dance, gymnastics) | 1-Never/seldom  2-1 day  3-2-3 days  4-4-5 days  5-6-7 days | | UB11 |
| **Self-perception Profile for Adolescents; Scale for Social Competence (adapted questions)** | | | |
| How well do the following statements correspond for you? | | | |
| 1. I find it quite hard to make friends | 1-Corresponds very poorly  2-Corresponds quite poorly  3-Corresponds quite well  4-Corresponds very well | | UB25 |
| 2. I have a lot of friends |  |  | UB26 |
| 3. Other teenagers find it hard to like me |  |  | UB27 |
| 4. I am popular with other teenagers |  |  | UB27 |
| 5. I feel socially accepted among others |  |  | UB28 |
| **Strengths and Difficulties Questionnaire (SDQ) – Prosocial Subscale** | | | |
| Give answers on the basis of your behaviour over the past 6 months. | | | |
| 1. I am considerate to other people’s feelings | 1- Not true  2- Somewhat true  3- Certainly true | | UB31 |
| 2. I share readily with others (treats, games other things) |  |  | UB32 |
| 3. I am helpful if someone is hurt, upset or feeling ill |  |  | UB33 |
| 4. I am kind to children younger than me |  |  | UB34 |
| 5. I often volunteer to help others (parents, teachers, other children/youths) |  |  | UB35 |
| **The Satisfaction with Life Scale (SWLS)** | | | |
| How satisfied are you with your life? | | | |
| 1. In most ways my life is close to my ideal | 1- Disagree completely  2- Disagree  3- Disagree somewhat  4- Don’t agree or disagree  5- Agree somewhat  6- Agree  7- Agree completely | | UB36 |
| 2. The conditions of my life are excellent |  |  | UB37 |
| 3. I am satisfied with my life |  |  | UB38 |
| 1. In most ways my life is close to my ideal 1- Disagree completely 4. So far I have gotten the important things I want in life |  |  | UB39 |
| 5. If I could live my life over, I would wish to have it the same  way |  |  | UB40 |
| **Short Mood and Feelings Questionnaire (SMFQ)** | | | |
| Here follows a list of different disturbing feelings and thoughts one might have sometimes. Think about the past two weeks and mark each item whether you have felt or thought these ways. | | | |
| 1. Felt miserable or unhappy | 1-Not true  2- Sometimes true  3-True | | UB41 |
| 2. Felt so tired that I just sat around and did nothing |  |  | UB42 |
| 3. Was very restless |  |  | UB43 |
| 4. Didn’t enjoy anything at all |  |  | UB44 |
| 5. Felt I was no good anymore |  |  | UB45 |
| 6. Cried a lot |  |  | UB46 |
| 7. Hated myself |  |  | UB47 |
| 8. Thought I could never be as good as other kids |  |  | UB48 |
| 9. Felt lonely |  |  | UB49 |
| 10. Thought nobody really loved me |  |  | UB50 |
| 11. Felt I was a bad person |  |  | UB51 |
| 12. Felt I did everything wrong |  |  | UB52 |
| 13. Found it hard to think/concentrate |  |  | UB53 |
| **The (Hopkins) Symptoms Checklist (SCL-10)** |  | |  |
| Have you over the past 2 weeks been bothered with any of the following? | | | |
| 1.Feeling fearful | 1-Not bothered  2-A little bothered  3-Quite bothered  4-Very bothered | | UB54 |
| 2.Nervousness or shakiness inside |  |  | UB55 |
| 3.Feeling hopeless about the future |  |  | UB56 |
| 4.Feeling blue |  |  | UB57 |
| 5.Worrying too much about things |  |  | UB58 |
| 6.Feeling everything is an effort |  |  | UB59 |
| 7.Feeling tense or keyed up |  |  | UB60 |
| 8.Suddenly scared for no reason |  |  | UB61 |
| 9.Anxiety or panic attack |  |  | UB62 |
| 10.Feelings of worthlessness |  |  | UB63 |
| **Mini Social Phobia Inventory (miniSPIN)** | | | |
| How much have the following problems bothered you during the past week? | | | |
| 1. Fear of embarrassment cause me to avoid doing things or speaking to people | 1-Not at all  2-A little bit  3-Somewhat  4-Very much  5-Extremely | | UB64 |
| 2. I avoid activities in which I am the centre of attention |  |  | UB65 |
| 3. Being embarrassed or looking stupid are among my worst fears |  |  | UB66 |
| **Scales concerning being bullied (BB) and bullying others (BO)** | | | |
| Bullying is defined as being excluded, teased, hit or bothered repeated times. Have you experienced being bullied over the past year? | | | |
| 1. Have been bullied by being teased | 1-Never  2-Now and then  3-Weekly  4-Daily | | UB67 |
| 2. Have been bullied by not being allowed to be with others, isolated or shut out from others |  |  | UB68 |
| 3. Have been bullied by being hit, kicked or pushed |  |  | UB69 |
| 4. Have been bullied by someone using mobile phones or other social media to spread rumours, tease or threaten you |  |  | UB70 |
| **Parental relations Self-concept Scale from the Self- Description Questionnaire II-Short (SDQII-S) (selective questions)** | | | |
| Here follows statements about how adolescents can feel about their parents. Mark each question how often you feel this way in your family | | | |
| 1. My parents understand me | 1-Never  2-Now and then  3-Often  4-Almost all the time | | UB71 |
| 2. I get along well with my parents |  |  | UB72 |
| 3. My parents like me |  |  | UB73 |
| 4. I like my parents |  |  | UB74 |
| **Parent-Child Conflict Scale; the Parental Environment Questionnaire (selected questions)** | | | |
| 5. My parents criticise me | 1-Never  2-Now and then  3-Often  4-Almost all the time | | UB75 |
| 6. My parents irritate me |  |  | UB76 |
| 7. My parents hurt my feelings |  |  | UB77 |
| 8. My parents and I get into arguments |  |  | UB78 |
| **Eating Disorder Examination Questionnaire (EDE-Q) (selected questions)** | | | |
| 1. When you think about the past 4 weeks, how often have you been deliberately trying to limit the amount of food you eat to influence your shape or weight? | 1-Never/rarely  2-Sometimes  3-Often  4-Very often | | UB79 |
| 2. Over the past 4 weeks, how often have you tried to follow definite rules regarding what you can eat, in order to influence your shape or weight (for example a limited amount of calories)? |  |  | UB80 |
| 3. Over the past 4 weeks, how often have you had a definite fear of losing control over eating? |  |  | UB81 |
| 4. Over the past 4 weeks, has thinking about food, eating or calories made it very difficult to concentrate on things you are interested in (for example, working, following a conversation, or reading)? |  |  | UB82 |
| 5. Over the past 4 weeks, have you eaten secretly? |  |  | UB83 |
| 6. How dissatisfied have you been with your shape (what you see in the mirror)? | 1-Not at all  2-A little  3-A lot  4-Very much | | UB84 |
| 7. How uncomfortable have you felt seeing your own body (for example seeing your shape in the mirror, while undressing, taking a bath or shower? |  |  | UB85 |
| 8. How uncomfortable have you felt about others seeing your shape or figure (for example in communal changing rooms, when swimming or wearing tight clothes)? |  |  | UB86 |
| **Sleeping problems** | | | |
| How often do you find it difficult to get to sleep at  night? | 1- Never  2- Less than once a week  3- Once per week  4- Twice per week  5- Three times per week  6- 4 times or more per week | | UB100 |
| How often have you woken up repeatedly during the night? |  |  | UB102 |
| How often do you feel tired or sleepy during the day? |  |  | UB104 |
| **Rating Scale for Disruptive Behaviour Disorders (RS-DBD)** | | | |
| Have you joined in or done any of this the past year? | | | |
| 1. Bullied, threatened or intimidated others | 1- Never/rarely  2- 1 time  3- 2-4 times  4- 5-10 times  5- 11-20 times  6- more than 20 times | | UB127 |
| 2. Initiated physical fights |  |  | UB128 |
| 3. Been physically cruel to others |  |  | UB129 |
| 4. Harassed or injured animals physically |  |  | UB130 |
| 5. Stolen items of nontrivial value without confronting a victim (e.g. shoplifting) |  |  | UB131 |
| 6. Deliberately destroyed other’s property |  |  | UB132 |
| 7. Been truant from school |  |  | UB133 |
| 8. Used an object that can cause serious physical harm to others (e.g. a bat, stone, knife, heavy toy) |  |  | UB134 |
| **The International Personality Item Pool (IPIP) Big-Five factor markers** | | | |
| Describe yourself the way you usually are | | | |
| *Extraversion* | | | |
| Describe yourself the way you usually are | | | |
| 1. Am the life of the party | 1-Strongly disagree  2-Disagree somewhat  3-Neither nor  4-Agree somewhat  5-Strongly agree | | UB135 |
| 6. Don’t talk a lot |  |  | UB140 |
| 11. Talk to a lot of different people at parties |  |  | UB145 |
| 16. Keep in the background |  |  | UB150 |
| *Agreeableness* |  |  |  |
| 2. Sympathize with others’ feelings |  |  | UB136 |
| 7. Am not interested in other people’s problems |  |  | UB141 |
| 12. Feel others’ emotions |  |  | UB146 |
| 17. Am not really interested in others |  |  | UB151 |
| *Conscientiousness* |  |  |  |
| 3. Get chores done right away |  |  | UB137 |
| 8. Often forget to put things back in their proper place |  |  | UB142 |
| 13. Like order |  |  | UB147 |
| 18. Make a mess of things |  |  | UB152 |
| *Emotional Stability* |  |  |  |
| 4. Have frequent mood swings |  |  | UB138 |
| 9. Am relaxed most of the time |  |  | UB143 |
| 14. Get upset easily |  |  | UB148 |
| 19. Often feel blue |  |  | UB153 |
| *Intellect* |  |  |  |
| 5. Have a vivid imagination |  |  | UB139 |
| 10. Am interested in abstract ideas |  |  | UB144 |
| 15. Have difficulty understanding abstract ideas |  |  | UB149 |
| 20. Do not have good imagination |  |  | UB154 |
| **Screen for Child Anxiety Related Disorders (SCARED)** | | | |
| Children and youth might be anxious at times, or be bothered by strange thoughts. Consider the past months and mark each item the way that best applies to you | | | |
| 1. I have been really frightened for no reason at all | 1-Not true  2-Sometimes true  3-Very true | | UB166 |
| 2. I have been afraid to be alone in the house |  |  | UB167 |
| 3. People have told me that I worry too much |  |  | UB168 |
| 4. I have been scared to go to school |  |  | UB169 |
| 5. I have been shy |  |  | UB170 |
| **Rosenberg Self Esteem Scale (RSES) (selective questions)** | | | |
| How do you feel about yourself? | | | |
| 1. I have a positive attitude toward myself | 1-Strongly disagree  2-Disagree  3-Agree  4-Strongly agree | | UB174 |
| 2. I feel completely useless at times |  |  | UB175 |
| 3. I feel that I do not have much to be proud about |  |  | UB176 |
| 4. I feel that I am a valuable person, as good as anyone else |  |  | UB177 |
| **Differential Emotional Scale (DES), Enjoyment Subscale** | | | |
| Think about the past two weeks. How often have you experienced this? | | | |
| 1. Felt glad about something | 1-Rarely or never  2-Hardly ever  3-Sometimes  4-Often  5-Very often | | UB178 |
| 2. Felt happy |  |  | UB179 |
| 3. Felt joyful, like everything was going your way |  |  | UB180 |
| **Questions about life events** | | | |
| Have you experienced any of these statements? Mark one or two boxes for each question. | | | |
|  | No | Yes, in last year | Yes, earlier |
| Have been seriously ill | UB181 | UB182 | UB183 |
| Have been involved in a serious accident | UB184 | UB185 | UB186 |
| Have been beaten, assaulted or badly humiliated | UB199 | UB200 | UB201 |
| Have lost someone close to you | UB211 | UB212 | UB213 |
| Have you any experienced mental health problems in the family | UB214 | UB215 | UB216 |
| Have you experienced suicide or suicide attempts in the family | UB217 | UB218 | UB219 |
| **Smoking habits** | | | |
| Do you smoke? | 1-Have never smoked  2-Have tried  3-Smoke now and then  4-Smoke daily | | UB239 |
| Do you use “snus”? | 1-Have never tried “snus”  2-Have tried  3-Use “snus” and then  4-Use “snus” daily | | UB242 |
| Do you use any of the following? |  | |  |
| E-cigarettes with nicotine | 1-Never  2-Have tried  3-Occasionally  4-Ddaily | | UB244 |
| Nicotine chewing gum |  |  | UB245 |
| Other nicotine preparations |  |  | UB246 |
| **Alcohol and drugs** | | | |
| Have you ever been drinking alcohol (more than just a sip) | 1-No  2-Yes | | UB247 |
| Drunk so much alcohol that you have obviously been intoxicated (drunk)? | 1-No, never  2-Once  3-2-5 times  4-6-10 times  5-More than 10 times | | UB249 |
| Used hashish/weed or marijuana? |  |  | UB250 |
| Used other drugs to become intoxicated? |  |  | UB251 |
| **The Perceived Stress Scale (PSS-4)** | | | |
| How are you typically as a person? Over the past month, how often have you felt… | | | |
| … That you were unable to control the important things in life? | 1-Never  2-Almost never  3-Sometimes  4-Quite often  5-Very often | | UB284 |
| … Confident about your ability to handle your personal problems? |  |  | UB285 |
| … That you succeeded with everything? |  |  | UB286 |
| … Difficulties were piling up so high that you could not overcome them? |  |  | UB287 |

*The table provides a brief overview of the Q-14year instruments used in this study. Detailed information about all MoBa questionnaires, including descriptions of all instruments and variables, is available on the official website of the Norwegian Mother, Father, and Child Study:* [*https://www.fhi.no/op/studier/moba/*](https://www.fhi.no/op/studier/moba/)*.*

### Table S2. Descriptive statistics for predictors from the clinical and family history, health and lifestyle, mental health symptoms, and psychosocial domains.

| **Predictor name** | **Mean (SD) or N (%)** | | |
| --- | --- | --- | --- |
|  | **Full sample**  **(n = 13,743)** | **Training set**  **(n = 10,994)** | **Test set**  **(n = 2,749)** |
| *1. Sociodemographic factors* | | | |
| Age in years when the questionnaire was filled, mean (SD) | 14.45 (0.52) | 14.44 (0.52) | 14.46 (0.52) |
| Female sex assigned at birth, N (%) | 7,242 (52.70%) | 5,782 (52.59%) | 1,460 (53.11%) |
| Immigrant status, N (%) | 110 (0.80%) | 85 (0.77%) | 25 (0.91%) |
| Parental education low, N (%) | 2,199 (16.00%) | 1,752 (15.94%) | 447 (16.26%) |
| Household income low, N (%) | 197 (1.43%) | 157 (1.43%) | 40 (1.46%) |
| *2. Clinical and family history* | | | |
| Neurodevelopmental disorder diagnosis | 406 (2.95%) | 330 (3.00%) | 76 (2.76%) |
| Other childhood-onset psychiatric disorder diagnosis | 684 (4.98%) | 554 (5.04%) | 130 (4.73%) |
| Other ICD-10 F-diagnosis | 299 (2.18%) | 241 (2.19%) | 58 (2.11%) |
| Parental internalizing disorder diagnosis | 3,062 (22.28%) | 2,410 (21.92%) | 652 (23.72%) |
| Primary care psychological symptom codes (ICPC-2) | 438 (3.19%) | 357 (3.25%) | 81 (2.95%) |
| *3. Health and lifestyle factors* | | | |
| Sleep problems | 8.24 (3.67) | 8.26 (3.68) | 8.16 (3.65) |
| Alcohol use | 4.36 (1.00) | 4.36 (1.00) | 4.34 (0.96) |
| Smoking | 5.31 (0.97) | 5.31 (0.96) | 5.32 (1.03) |
| Exercise | 3.15 (1.20) | 3.14 (1.20) | 3.20 (1.20) |
| Time spent watching TV | 2.79 (0.98) | 2.79 (0.99) | 2.79 (0.97) |
| Time spent gaming | 2.58 (1.27) | 2.58 (1.28) | 2.56 (1.24) |
| Time spent using social media | 3.32 (1.16) | 3.32 (1.16) | 3.31 (1.16) |
| *4. Mental health symptoms* | | | |
| SMFQ (depressive symptoms) | 19.87 (5.77) | 19.89 (5.78) | 19.79 (5.73) |
| SCL-10 (psychological distress) | 16.62 (5.92) | 16.66 (5.94) | 16.47 (5.85) |
| Mini-SPIN (social anxiety) | 5.72 (2.88) | 5.72 (2.89) | 5.72 (2.84) |
| SCARED (anxiety symptoms) | 6.63 (1.78) | 6.63 (1.78) | 6.61 (1.77) |
| EDE-Q (eating disorder symptoms) | 11.98 (4.48) | 12.00 (4.51) | 11.90 (4.35) |
| RS-DBD (disruptive behaviour) | 8.74 (1.89) | 8.76 (1.91) | 8.70 (1.79) |
| *5. Psychosocial domain* | | | |
| SPA – Scale for Social Competence | 16.02 (2.75) | 16.01 (2.74) | 16.03 (2.79) |
| SDQ – Prosocial Subscale | 13.30 (1.59) | 13.29 (1.59) | 13.30 (1.61) |
| Satisfaction with Life Scale | 27.32 (6.20) | 27.32 (6.17) | 27.32 (6.33) |
| DES – Enjoyment Subscale | 4.13 (0.80) | 4.13 (0.81) | 4.16 (0.79) |
| SDQII-S – Relation with Parents | 14.20 (2.08) | 14.19 (2.08) | 14.25 (2.09) |
| PEQ – The Parent-Child Conflict Scale | 6.88 (1.90) | 6.89 (1.91) | 6.87 (1.88) |
| Bullying scale | 4.45 (0.98) | 4.46 (0.98) | 4.43 (0.96) |
| Rosenberg Self-Esteem Scale | 12.28 (2.66) | 12.28 (2.67) | 12.30 (2.64) |
| IPIP Big-Five Factors - extraversion | 13.10 (3.79) | 13.10 (3.80) | 13.13 (3.73) |
| IPIP Big-Five Factors - conscientiousness | 14.21 (2.92) | 14.22 (2.93) | 14.17 (2.86) |
| IPIP Big-Five Factors - emotional stability | 13.36 (3.39) | 13.35 (3.39) | 13.39 (3.40) |
| IPIP Big-Five Factors - intellect | 14.28 (2.90) | 14.29 (2.90) | 14.25 (2.90) |
| IPIP Big-Five Factors - agreeableness | 16.27 (2.58) | 16.27 (2.58) | 16.26 (2.59) |
| Serious Life Event | 7,094 (51.62%) | 5,699 (51.84%) | 1,395 (50.75%) |

Values are presented as mean (SD) for continuous variables and N (%) for categorical variables.

### Table S3. Stratifying test-set participants by predicted risk of internalizing disorder

| **Model** | **OR80/20 (95% CI)** | **ORmid/20 (95% CI)** | **OR80/rest (95% CI)** |
| --- | --- | --- | --- |
| Model 1 | 3.42 (1.96-5.97) | 2.61 (1.52-4.58) | 1.64 (1.19-2.16) |
| Model 2 | 5.15 (3.03-8.46) | 3.28 (1.98-5.46) | 2.07 (1.49-2.82) |
| Model 3 | 18.06 (7.36-48.00) | 7.01 (2.93-19.79) | 3.31 (2.40-4.32) |
| Model 4 | 19.77 (8.00-50.68) | 7.67 (3.27-19.83) | 3.32 (2.39-4.48) |
| Model 5 | 13.85 (6.68-30.35) | 4.67 (2.18-10.79) | 3.73 (2.75-5.01) |
| Model 6 | 17.02 (7.41-38.94) | 5.41 (2.45-12.40) | 3.98 (2.87-5.38) |

OR80/20: odds ratios comparing high-risk group (≥80^th^ percentile) and low-risk group (≤20^th^ percentile); ORmid/20: odds ratios comparing medium-risk group (21-79^th^ percentile) and low-risk group; OR80/rest: odds ratios comparing high-risk group with the rest of the sample.

Model 1: sociodemographic factors; Model 2: model 1 + clinical and family history; Model 3: model 2 + health and lifestyle factors; Model 4: model 3 + mental health symptoms; Model 5: model 4 + psychosocial domain; Model 6 (full model): model 5 + polygenic scores.

Values are presented as estimates with 95% confidence intervals (CIs) obtained by 1,000 bootstrap resamples.

### Table S4. Predictive performance of individual predictor groups

| **Predictor sets** | | **AUC test (95% CI)** | **∆AUC (95% CI)** |
| --- | --- | --- | --- |
| Set 0 | Age, sex | 0.610 (0.577-0.644) | NA |
| Set 1 | Set 0 + other sociodemographic factors | 0.626 (0.588-0.663) | 0.0159 (0.00-0.033) |
| Set 2 | Set 0 + clinical and family history | 0.639 (0.605-0.676) | 0.0282 (0.0046-0.0506) |
| Set 3 | Set 0 + lifestyle factors | 0.699 (0.660-0.733) | 0.0884 (0.0582-0.124) |
| Set 4 | Set 0 + mental health symptoms | 0.710 (0.671-0.743) | 0.1000 (0.0632-0.138) |
| Set 5 | Set 0 + psychosocial domain | 0.703 (0.664-0.737) | 0.0934 (0.0601-0.128) |
| Set 6 | Set 0 + polygenic scores | 0.666 (0.626-0.703) | 0.0549 (0.0262-0.0804) |

AUC: area under the receiver operating characteristic curve; CI: confidence interval; NA: not applicable. Test AUC values are presented as estimates with 95% confidence intervals (CIs) obtained by 1,000 bootstrap resamples.

The change in AUC (∆AUC) was calculated as the difference between each model (Sets 1-6) and the baseline model (Set 0), and both ∆AUC and its 95% CIs were estimated using 1,000 bootstrap resamples.

### Table S5. Performance metrics of the nested models in the test set at the thresholds determined in the training set.

**A) Performance metrics in the test set, estimated at the optimal threshold determined by Youden’s index in the training set.**

| **Model** | **Specificity (95% CI)** | **Sensitivity (95% CI)** | **Balanced accuracy (95% CI)** | **PPV (95% CI)** | **NPV (95% CI)** | **Threshold** |
| --- | --- | --- | --- | --- | --- | --- |
| 1 | 0.486 (0.467-0.505) | 0.750 (0.692-0.811) | 0.619 (0.587-0.649) | 0.103 (0.095-0.111) | 0.961 (0.952-0.970) | 0.079 |
| 2 | 0.474 (0.454-0.493) | 0.781 (0.726-0.831) | 0.627 (0.599-0.656) | 0.105 (0.098-0.112) | 0.965 (0.956-0.973) | 0.079 |
| 3 | 0.557 (0.537-0.577) | 0.710 (0.652-0.771) | 0.634 (0.600-0.665) | 0.112 (0.103-0.122) | 0.961 (0.952-0.969) | 0.060 |
| 4 | 0.732 (0.715-0.750) | 0.557 (0.493-0.627) | 0.645 (0.611-0.680) | 0.141 (0.125-0.159) | 0.954 (0.948-0.962) | 0.076 |
| 5 | 0.648 (0.630-0.667) | 0.677 (0.612-0.741) | 0.662 (0.628-0.695) | 0.132 (0.119-0.144) | 0.962 (0.954-0.970) | 0.062 |
| 6 | 0.701 (0.682-0.717) | 0.632 (0.562-0.701) | 0.666 (0.629-0.702) | 0.143 (0.128-0.157) | 0.960 (0.953-0.968) | 0.072 |

**B) Performance metrics estimated in the test set, estimated at the threshold selected in the training set to target 50% sensitivity.**

| **Model** | **Specificity (95% CI)** | **Sensitivity (95% CI)** | **Balanced accuracy (95% CI)** | **PPV (95% CI)** | **NPV (95% CI)** | **Threshold** |
| --- | --- | --- | --- | --- | --- | --- |
| 1 | 0.726 (0.709-0.743) | 0.378 (0.313-0.453) | 0.552 (0.518-0.593) | 0.0983 (0.082-0.118) | 0.937 (0.930-0.944) | 0.0954 |
| 2 | 0.651 (0.633-0.669) | 0.498 (0.438-0.567) | 0.575 (0.542-0.610) | 0.101 (0.089-0.114) | 0.943 (0.936-0.950) | 0.0849 |
| 3 | 0.793 (0.778-0.808) | 0.438 (0.368-0.508) | 0.615 (0.581-0.651) | 0.143 (0.122-0.165) | 0.947 (0.941-0.954) | 0.0988 |
| 4 | 0.832 (0.816-0.847) | 0.403 (0.343-0.473) | 0.617 (0.586-0.652) | 0.159 (0.135-0.184) | 0.946 (0.941-0.952) | 0.102 |
| 5 | 0.843 (0.830-0.857) | 0.428 (0.363-0.493) | 0.636 (0.600-0.670) | 0.177 (0.150-0.204) | 0.949 (0.943-0.955) | 0.108 |
| 6 | 0.836 (0.821-0.849) | 0.438 (0.373-0.512) | 0.637 (0.602-0.673) | 0.174 (0.147-0.200) | 0.950 (0.944-0.956) | 0.107 |

In table A), each model was assessed at its own threshold fixed from the training set, resulting in different operating points across models. In table B), the threshold for each model was also fixed from the training set; consequently, the achieved test-set sensitivity may deviate from 50%.

PPV: positive predictive value; NPV: negative predictive value. Model 1: sociodemographic factors; Model 2: model 1 + clinical and family history; Model 3: model 2 + health and lifestyle factors; Model 4: model 3 + mental health symptoms; Model 5: model 4 + psychosocial domain; Model 6 (full model): model 5 + polygenic scores.

Values are presented as estimates with 95% confidence intervals (CIs) obtained by 1,000 bootstrap resamples.

### Table S6. Performance of the full model in leave-one-region-out validation

| **Region used as a hold-out test set** | **Size of the training set** | **AUC test**  **(95% CI)** |
| --- | --- | --- |
| Region 2 (n = 3,510) | n = 10,233 | 0.761 (0.732-0.788) |
| Region 3 (n = 1,802) | n = 11,941 | 0.762 (0.722-0.806) |
| Region 4 (n = 833) | n = 12,910 | 0.750 (0.686-0.810) |
| Region 1 (n = 7,598) | n = 6,145 | 0.749 (0.727-0.769) |

AUC: area under the receiver operating characteristic curve. Values are presented as estimates with 95% confidence intervals (CIs) obtained by 1,000 bootstrap resamples.

Region 1: South-Eastern Norway (Helse Sør-Øst RHF); Region 2: Western Norway (Helse Vest RHF); Region 3: Central Norway (Helse Midt-Norge RHF); Region 4: Northern Norway (Helse Nord RHF).

### Table S7. Performance of the full model across different alpha values in the test set.

| **Alpha** | **AUC (95% CI)** | **Specificity (95% CI)** | **Sensitivity (95% CI)** | **Balanced accuracy (95% CI)** | **PPV**  **(95% CI)** | **NPV**  **(95% CI)** |
| --- | --- | --- | --- | --- | --- | --- |
| 0.00 | 0.730  (0.696-0.765) | 0.721  (0.704-0.738) | 0.606  (0.537-0.677) | 0.664  (0.627-0.699) | 0.146  (0.131-0.163) | 0.959  (0.951-0.966) |
| 0.25 | 0.732  (0.697-0.764) | 0.707  (0.690-0.724) | 0.622  (0.557-0.692) | 0.664  (0.630-0.700) | 0.143  (0.128-0.158) | 0.960  (0.953-0.967) |
| 0.75 | 0.732  (0.697-0.766) | 0.702  (0.686-0.719) | 0.628  (0.562-0.692) | 0.665  (0.629-0.700) | 0.143  (0.128-0.158) | 0.960  (0.953-0.967) |
| 1.00 | 0.732  (0.695-0.766) | 0.642  (0.623-0.661) | 0.691  (0.627-0.751) | 0.667  (0.632-0.699) | 0.132  (0.120-0.144) | 0.963  (0.956-0.971) |

Performance metrics were estimated at the optimal threshold determined by Youden’s index. PPV: positive predictive value; NPV: negative predictive value; AUC: area under the receiver operating characteristic curve. Values are presented as estimates with 95% confidence intervals (CIs) obtained by 1,000 bootstrap resamples.

### Table S8. Stratifying test-set participants by predicted risk of internalizing disorder using selected triplet models.

| **Triplet** | **Predictors** | **OR80/20**  **(95% CI)** | **ORmid/20 (95% CI)** | **OR80/rest (95% CI)** |
| --- | --- | --- | --- | --- |
| 1 | SCL-10 + Sleep + Exercise | 9.36 (4.68-18.55) | 3.30 (1.72-6.76) | 3.46 (2.45-4.61) |
| 2 | SPA-SSC + IPIP_stab + SWLC | 8.91 (4.78-17.27) | 3.31 (1.63-6.10) | 3.51 (2.49-4.73) |
| 3 | SCARED + RSES + DES | 8.41 (4.54-16.70) | 3.24 (1.76-6.43) | 3.18 (2.32-4.27) |
| 4 | EDE-Q + SCL-10 + Exercise | 7.44 (4.05-13.61) | 2.49 (1.39-4.64) | 3.54 (2.58-4.72) |
| 5 | SCARED + SWLC + IPIP_stab | 6.39 (3.67-11.17) | 2.34 (1.37-4.07) | 3.21 (2.36-4.28) |
| 6 | Sleep + SMFQ + SPA-SSC | 10.60 (5.41-20.24) | 3.63 (1.92-7.12) | 3.60 (2.63-4.72) |
| 7 | EDE-Q + DES + SMFQ | 7.04 (3.87-12.97) | 2.78 (1.54-5.11) | 3.03 (2.32-4.13) |

OR80/20: odds ratios comparing high-risk group (≥80^th^ percentile) and low-risk group (≤20^th^ percentile); ORmid/20: odds ratios comparing medium-risk group (21-79^th^ percentile) and low-risk group; OR80/rest: odds ratios comparing high-risk group with the rest of the sample.

AUC: Area under the receiver operating characteristic curve; SCL-10: The (Hopkins) Symptoms Checklist; SPA-SSC: Self-Perception Profile for Adolescents, Scale for Social Competence (adapted questions); SWLC: Satisfaction with Life Scale; IPIP_stab: the International Personality Item Pool Big-Five factor markers, selective items related to emotional stability factor; SCARED: Screen for Child Anxiety Related Disorders; RSES: Rosenberg Self-Esteem Scale (selected questions); EDE-Q: Eating Disorder Examination Questionnaire (selected questions); DES: Differential Emotional Scale, Enjoyment Subscale; SMFQ: Short Mood and Feelings Questionnaire; Sleep: questions about sleeping problems.

95% confidence intervals (CIs) obtained by 1,000 bootstrap resamples.

### Supplementary Figures

### Figure S1. Flow diagram of the study sample.

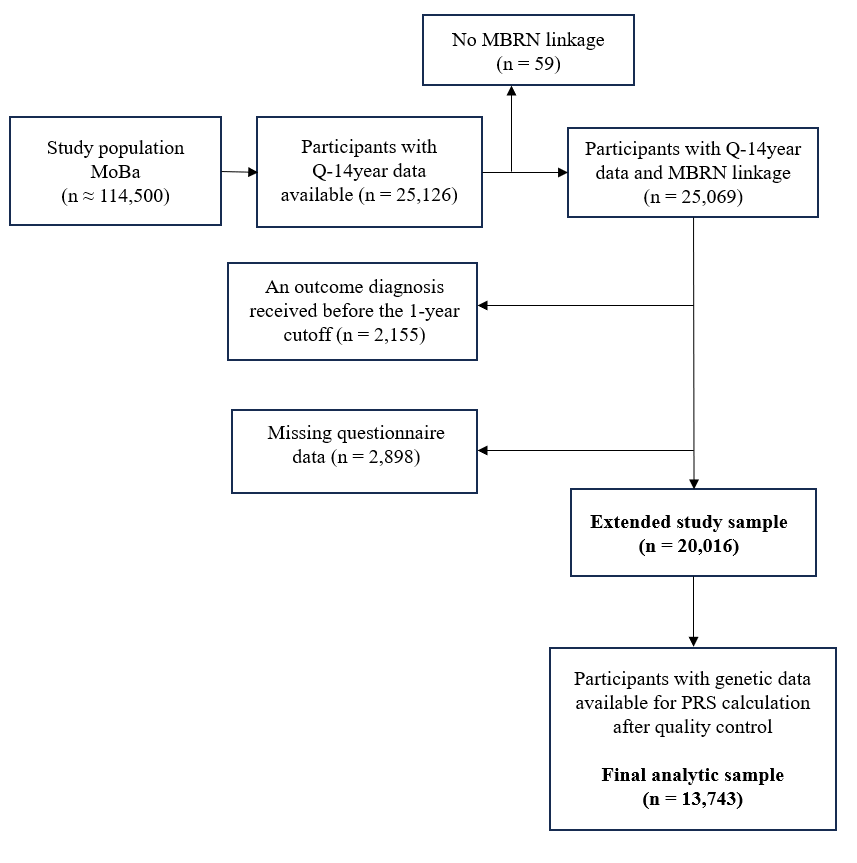

### Figure S2. Predictive performance of nested models in the test set.

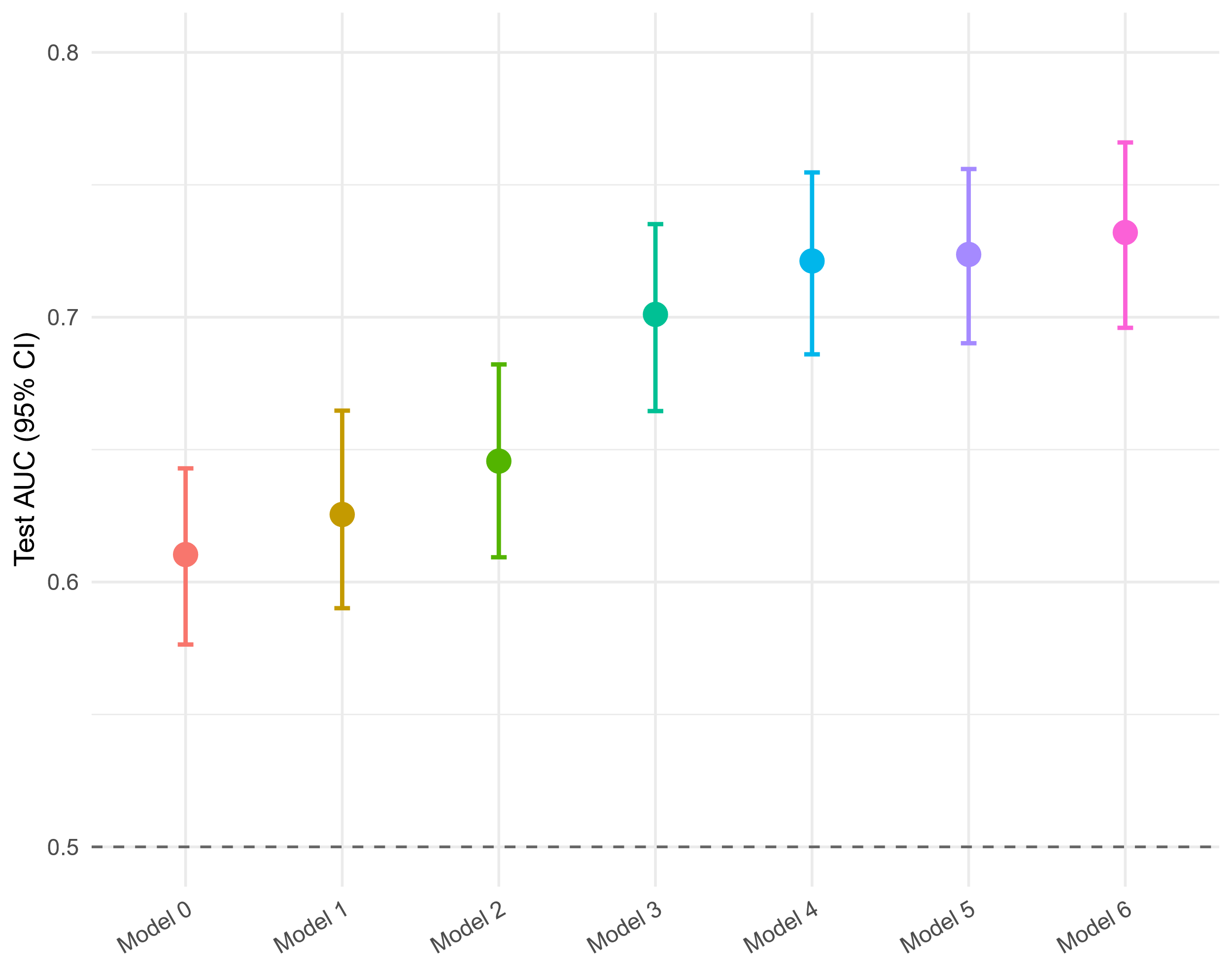

Test set predictive performance of nested models. Each point shows the test set area under the receiver operating characteristic curve (AUC); error bars represent 95% confidence intervals (CIs) obtained by 1,000 bootstrap resamples. Models include successively larger predictor sets. Model 1: sociodemographic factors; Model 2: model 1 + clinical and family history; Model 3: model 2 + health and lifestyle factors; Model 4: model 3 + mental health symptoms; Model 5: model 4 + psychosocial domain; Model 6 (full model): model 5 + polygenic scores.

### Figure S3. Predictive performance of individual predictor groups in the test set.

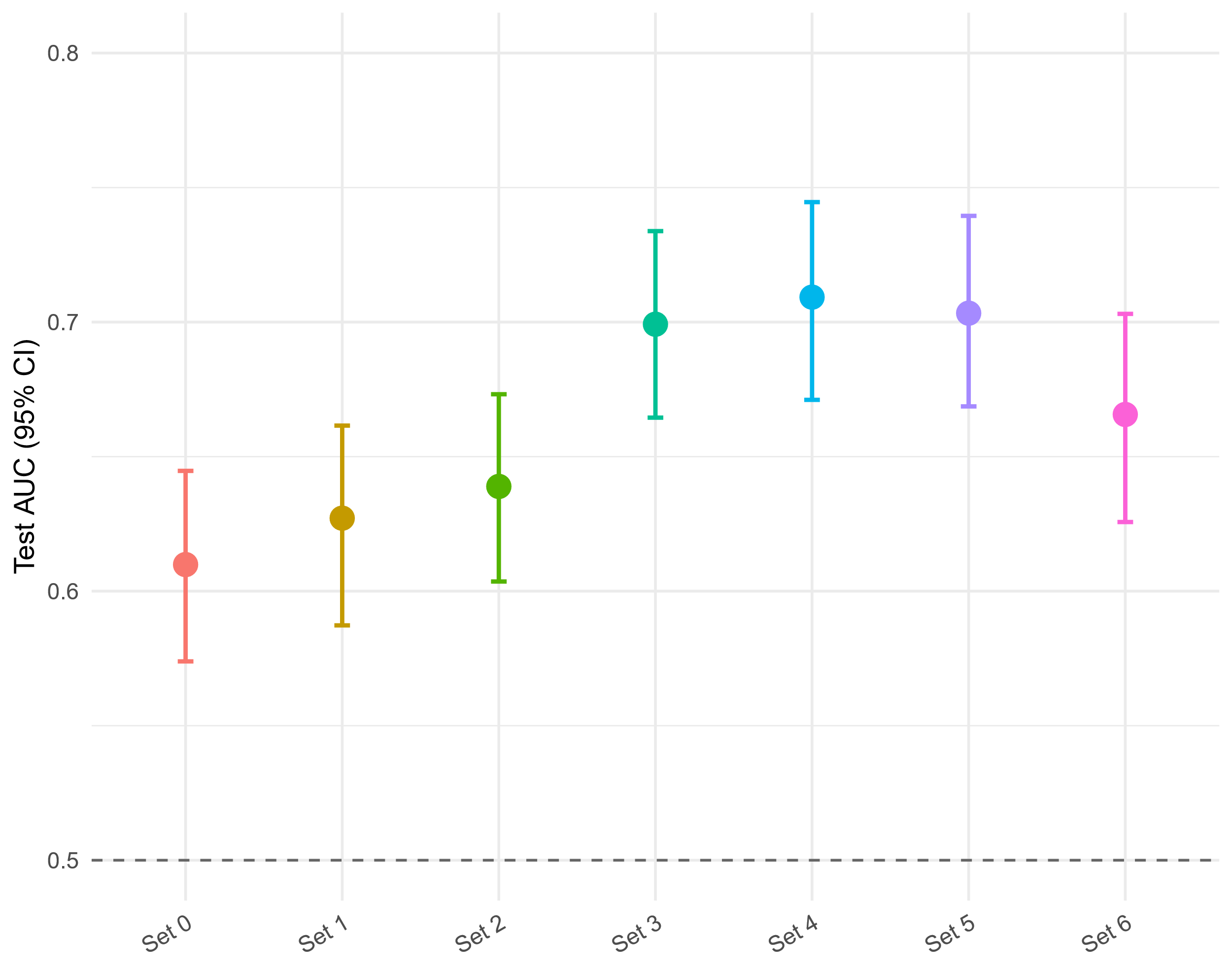

Predictive performance of models trained with individual predictor groups. Each point shows the test set area under the receiver operating characteristic curve (AUC); error bars represent 95% confidence intervals (CIs) obtained by 1,000 bootstrap resamples. Predictor sets were modelled separately, with age and sex included as baseline covariates. Set 0 = age, sex; set 1 = set 0 + other sociodemographic factors; set 2 = set 0 + clinical and family history; set 3 = set 0 + health and lifestyle factors; set 4 = set 0 + mental health symptoms; set 5 = set 0 + psychosocial domain; set 6 = set 0 + polygenic scores.

### Figure S4. Precision-recall curves of the nested models in the test set.

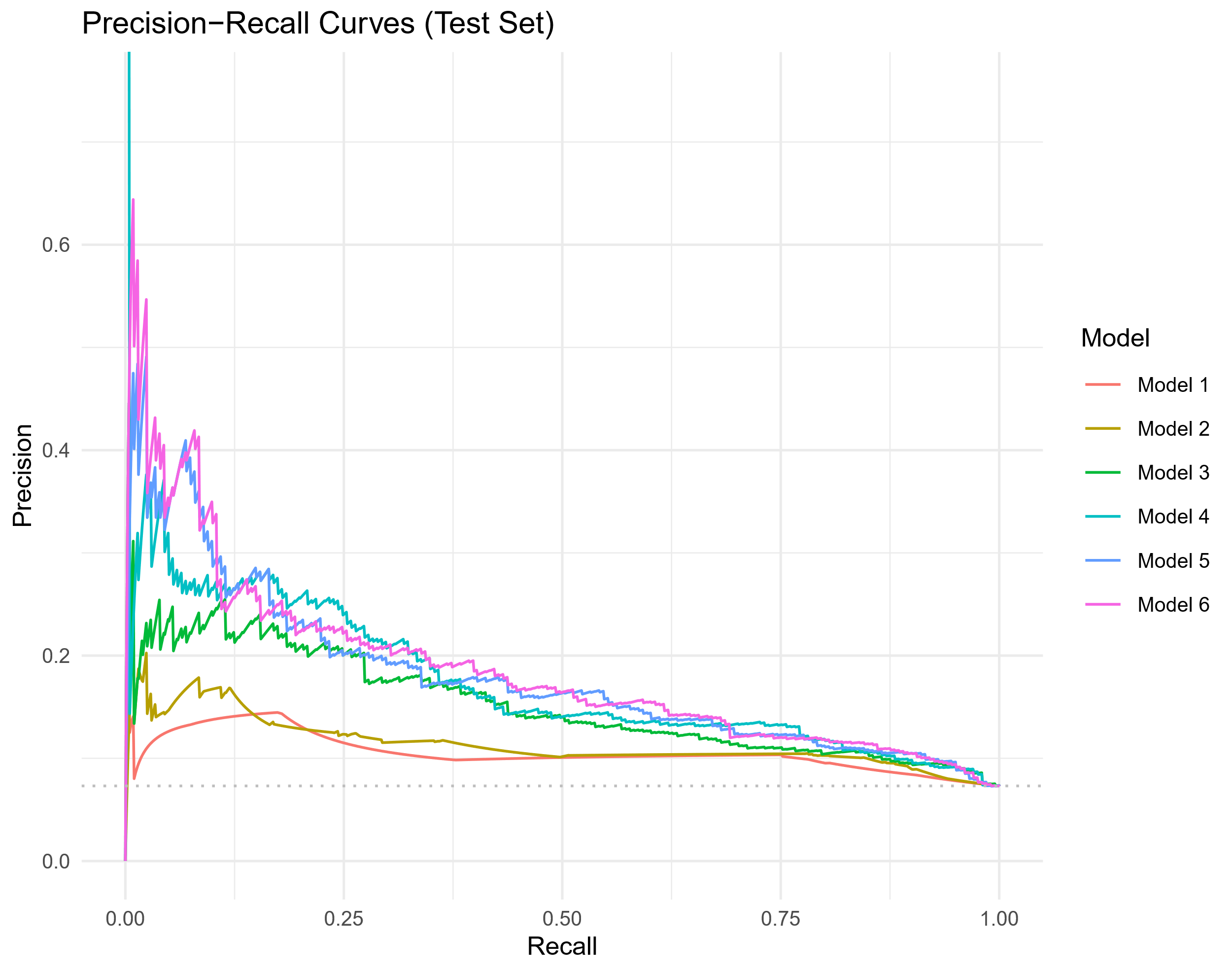

Models include successively larger predictor sets. Model 1: sociodemographic factors; Model 2: model 1 + clinical and family history; Model 3: model 2 + health and lifestyle factors; Model 4: model 3 + mental health symptoms; Model 5: model 4 + psychosocial domain; Model 6 (full model): model 5 + polygenic scores.

### Figure S5. Confusion matrices for nested models in the test set at thresholds determined in the training set.

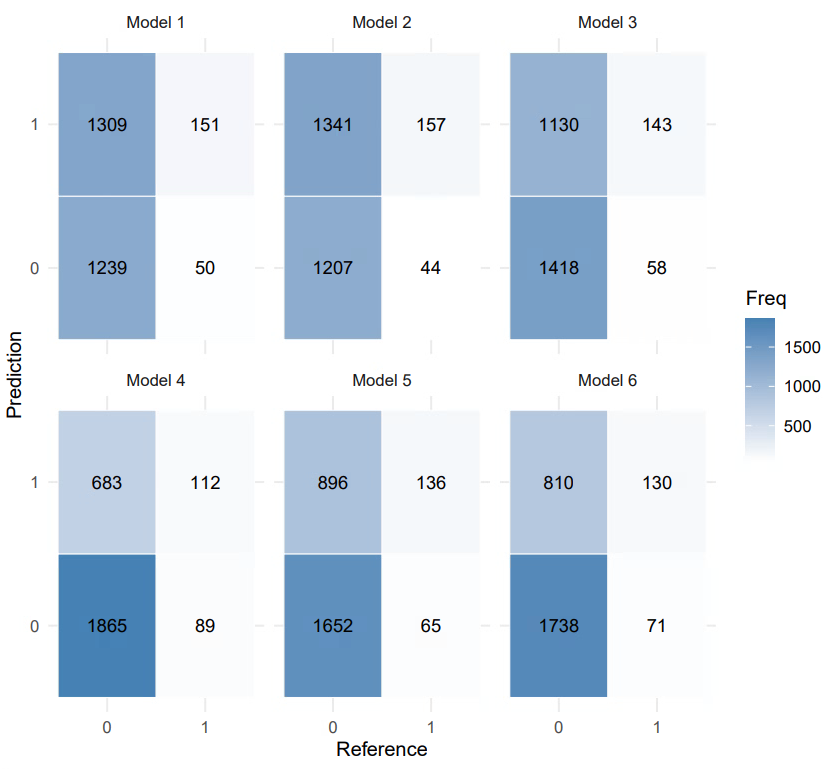

Confusion matrix for each model was calculated at the optimal threshold determined by Youden’s index in the training set. Cell counts indicate the number of participants in each category. Model 1: sociodemographic factors; Model 2: model 1 + clinical and family history; Model 3: model 2 + lifestyle factors; Model 4: model 3 + mental health symptoms; Model 5: model 4 + psychosocial domain; Model 6 (full model): model 5 + polygenic scores.

### Figure S6. Training performance of single and pairwise predictors (age/sex-adjusted) across health/lifestyle, mental health symptoms, and psychosocial domains.

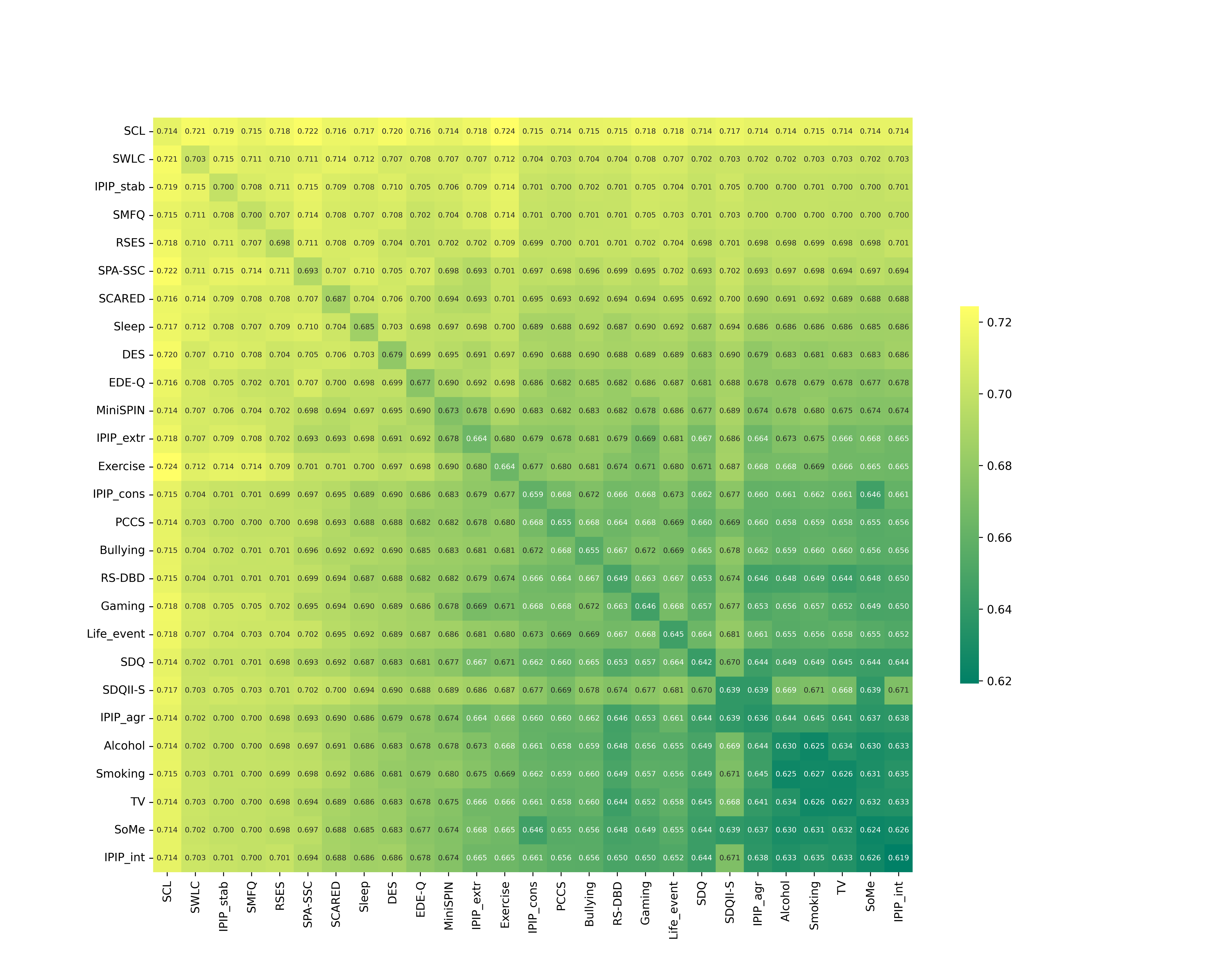

Heatmap showing area under the receiver operating characteristic curve (AUC) in the training set for models including age, sex, and one or two predictors from health and lifestyle factors, mental health symptoms, and the psychosocial domain. The diagonal represents models with age, sex, and a single predictor; off-diagonal cells represent models with predictor pairs.

SCL-10: The (Hopkins) Symptoms Checklist; SWLC: Satisfaction with Life Scale; IPIP_stab: the International Personality Item Pool Big-Five factor markers, selective items related to emotional stability factor; SMFQ: Short Mood and Feelings Questionnaire; SPA-SSC: Self-Perception Profile for Adolescents, Scale for Social Competence (adapted questions); RSES: Rosenberg Self-Esteem Scale (selected questions); SCARED: Screen for Child Anxiety Related Disorders; Sleep: questions about sleeping problems; DES: Differential Emotional Scale, Enjoyment Subscale; EDE-Q: Eating Disorder Examination Questionnaire (selected questions); MiniSPIN: Mini Social Phobia Inventory; IPIP_extr: the International Personality Item Pool Big-Five factor markers, selective items related to extraversion factor; Exercise: exercise, days per week; IPIP_cons: the International Personality Item Pool Big-Five factor markers, selective items related to conscientiousness factor; PCCS: Parent-Child Conflict Scale; Bullying: questions concerning being bullied; RS_DBD: Parent/Teacher Rating Scale for Disruptive Behaviour Disorders (selected questions); Gaming: time spent gaming; Life_event: serious life event; SDQ: Strengths and Difficulties Questionnaire, Prosocial Subscale; SDQII-S: Parental relations Self-concept Scale from the Self- Description Questionnaire II-Short (selected questions); IPIP_agr: the International Personality Item Pool Big-Five factor markers, selective items related to agreeableness factor; Alcohol: questions about experience with alcohol and use of drugs; Smoking: questions about the child’s smoking/snusing habits; TV: time spent watching TV; SoMe: time spent communicating with friends on social media; IPIP_int: the International Personality Item Pool Big-Five factor markers, selective items related to intellect factor.

### Figure S7. Training performance of triplet models (age/sex-adjusted) across health/lifestyle, mental health symptoms, and psychosocial domains.

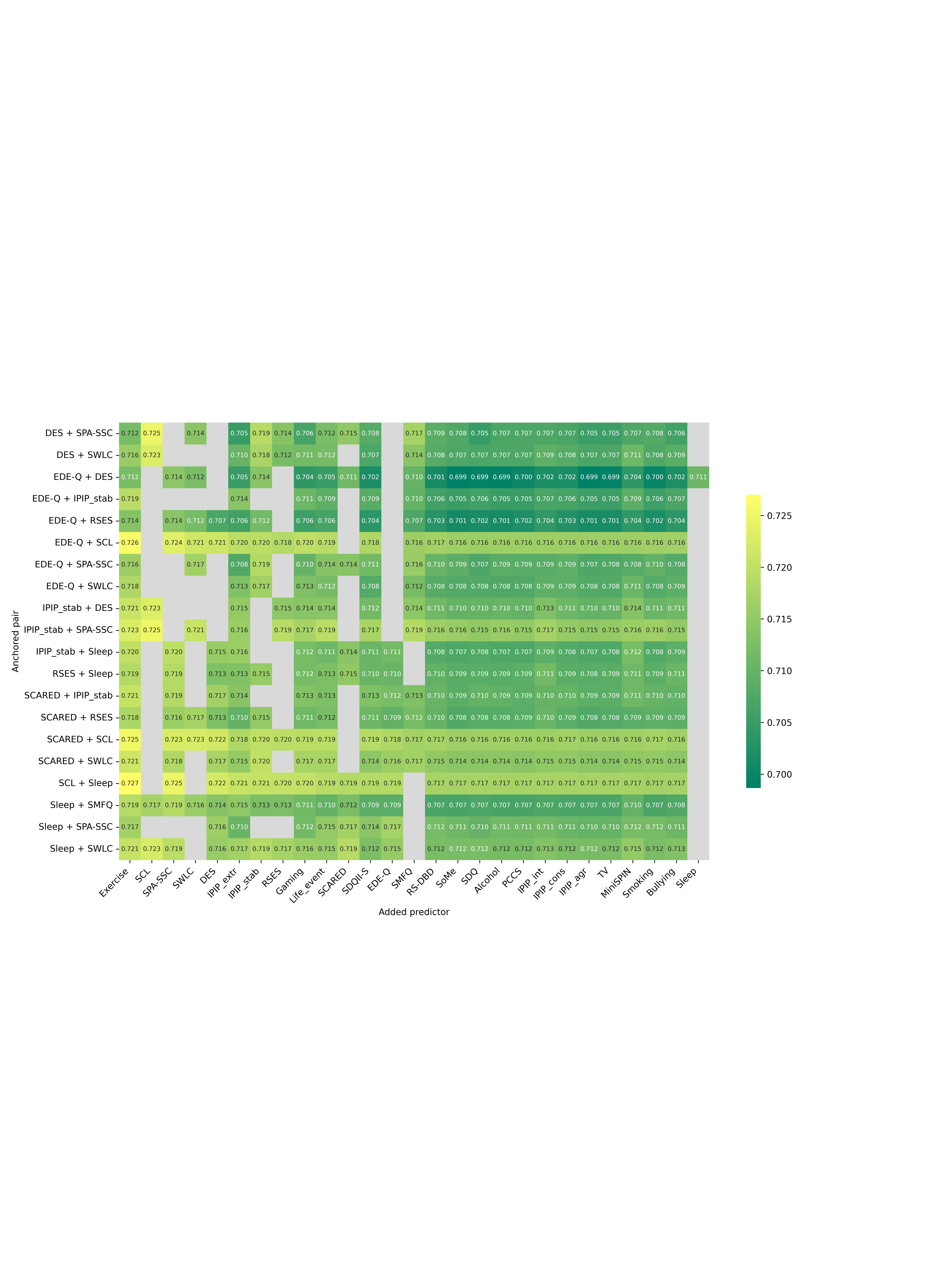

Heatmap showing area under the receiver operating characteristic curve (AUC) in the training set for models including age, sex, and three predictors from health and lifestyle factors, mental health symptoms, and the psychosocial domain. Rows show the anchored pair; columns show the added predictor. Grey tiles mark combinations not evaluated (i.e. either the added predictor duplicates a member of the pair, or the resulting triplet is a duplicate of an already assessed triplet).

SCL-10: The (Hopkins) Symptoms Checklist; SWLC: Satisfaction with Life Scale; IPIP_stab: the International Personality Item Pool Big-Five factor markers, selective items related to emotional stability factor; SMFQ: Short Mood and Feelings Questionnaire; SPA-SSC: Self-Perception Profile for Adolescents, Scale for Social Competence (adapted questions); RSES: Rosenberg Self-Esteem Scale (selected questions); SCARED: Screen for Child Anxiety Related Disorders; Sleep: questions about sleeping problems; DES: Differential Emotional Scale, Enjoyment Subscale; EDE-Q: Eating Disorder Examination Questionnaire (selected questions); MiniSPIN: Mini Social Phobia Inventory; IPIP_extr: the International Personality Item Pool Big-Five factor markers, selective items related to extraversion factor; Exercise: exercise, days per week; IPIP_cons: the International Personality Item Pool Big-Five factor markers, selective items related to conscientiousness factor; PCCS: Parent-Child Conflict Scale; Bullying: questions concerning being bullied; RS_DBD: Parent/Teacher Rating Scale for Disruptive Behaviour Disorders (selected questions); Gaming: time spent gaming; Life_event: serious life event; SDQ: Strengths and Difficulties Questionnaire, Prosocial Subscale; SDQII-S: Parental relations Self-concept Scale from the Self- Description Questionnaire II-Short (selected questions); IPIP_agr: the International Personality Item Pool Big-Five factor markers, selective items related to agreeableness factor; Alcohol: questions about experience with alcohol and use of drugs; Smoking: questions about the child’s smoking/snusing habits; TV: time spent watching TV; SoMe: time spent communicating with friends on social media; IPIP_int: the International Personality Item Pool Big-Five factor markers, selective items related to intellect factor.

### Figure S8. Training performance of triplets vs. pairs of predictors (age/sex-adjusted).

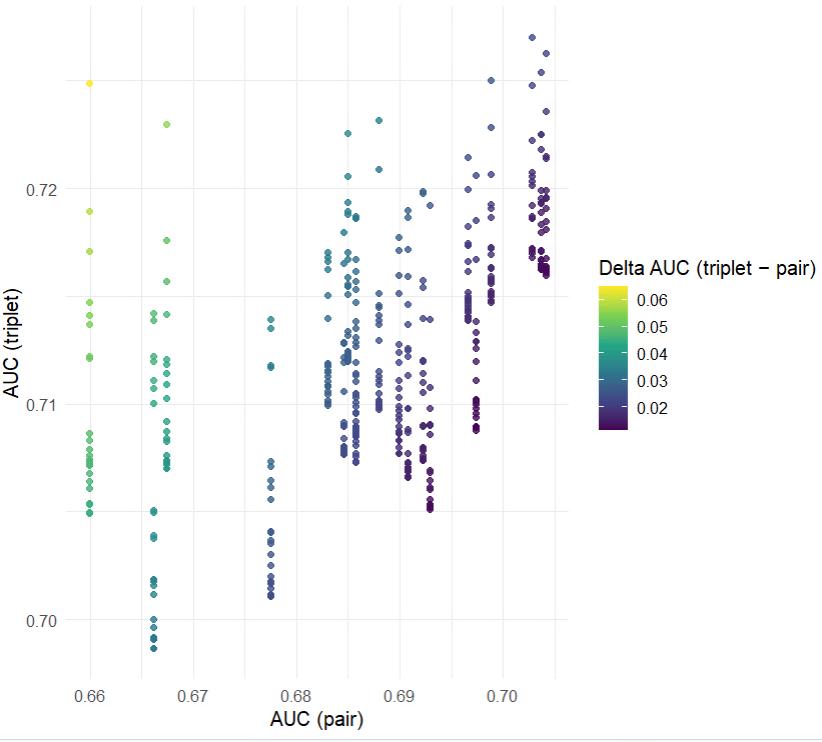

Scatterplot comparing each triplet’s performance with its anchored pair. The x-axis shows AUC_pair and the y-axis AUC_triplet; points are colored by ΔAUC = AUC_triplet − AUC_pair.
